# Effects of wearing FFP2 masks on SARS-CoV-2 infection rates in classrooms

**DOI:** 10.1101/2022.07.24.22277968

**Authors:** Gerald Jarnig, Reinhold Kerbl, Mireille N.M. van Poppel

## Abstract

**Importance:** Different mitigation measures are mandated in schools worldwide to control the spread of SARS-CoV-2. The efficacy of most measures, however, has not been investigated thus far.

**Objective:** To investigate the usefulness of FFP-2 masks in classrooms to prevent the spread of SARS-CoV-2.

**Design:** A retrospective comparative cohort study of infection rates (evaluated by PCR screening in school) in students wearing FFP-2 masks continuously and students in sports classes with limited face mask use.

**Setting:** A single-center evaluation comparing classes (middle school: age 10-16 years, 4-year high school: age 14-20 years) with a high sports focus (SF), with regular classes during the Delta and Omicron waves (September 2021–April 2022).

**Participants:** In total, 616 children/families were invited to participate in the comparative evaluation, and 614 (99.7%) followed this invitation by providing relevant information concerning their SARS-CoV-2 infection status. A total of 213 legal guardians (for children < 14 years) and 401 adolescents (≥14 years) reported SARS-CoV-2 infections during the 2021/22 school year.

**Main Outcomes and Measures:** A comparative analysis of cumulative SARS-CoV-2 infection rates in sports and non-sports classes (the 7-day classroom incidence of SARS-CoV-2 infections, and potential secondary infections among school classmates).

**Results:** Cumulative SARS-CoV-2 infection rates were clearly higher in sports classes (with limited mask use) than in non-sports classes (continuous mask use). After the relaxation of the mitigation measures, students in non-sports classes, however, showed a clear “catch-up” of infections, leading to a higher incidence of infections during this phase. By the end of the observation period (April 30, 2022), only a small difference in cumulative SARS-CoV-2 infection rates (p=0.037, φ=0.09) was detected between classes with a sports focus and those without a sports focus.

**Conclusions and Relevance:** Wearing FFP2 face masks reduces the risk of SARS-CoV-2 infection if strict mitigation measures are applied. Following the relaxation of strict measures, previously “protected” students show a significant “catch-up” infection rate. Thus, continuous face mask use postpones rather than avoids SARS-CoV-2 infection in many cases. Therefore, the advantage of reduced transmission must be carefully balanced against the disadvantages associated with mask wearing throughout schools.

## INTRODUCTION

Worldwide, different restrictions were mandated to control the spread of different severe acute respiratory syndrome coronavirus 2 (SARS-CoV-2) variants. The stringency of these was based on the predominant viral load in the national population.^1^ As one of these measures, the usefulness and practicality of medical face masks to contain the spread of viruses has been proven.^2,3^ Increasing numbers of studies are reporting the usefulness of FFP2 masks to reduce the spread of SARS-CoV-2.^4-6^ In public spaces, in addition to strict hygiene rules and a minimum safety distance, the use of face masks has been recommended as they have been shown to greatly reduce the spread of different virus variants.^7^

In June 2020, a recommendation was published by the World Health Organization (WHO) describing the use of face masks (mouth–nose masks) in the context of coronavirus disease 2019 (COVID-19).^8^ On August 21, 2020, this recommendation was expanded to detail describing how children should use a face mask in the community.^9^

Thus far, no negative physical effects of face masks have been proven for children.^10-12^ However, an increasing number of studies report psychological problems triggered by continued mandatory use of these.^13-15^ Therefore, it appears essential to carefully balance the advance of reduced virus transmission against the negative behavioral and psychological aspects, as well as in the light of concerns about the correct wearing of face masks by children over a long period.^16-18^.

The aim of this study was to assess if wearing face masks in the classroom is a useful mitigation measure to control SARS-CoV-2 infections and to gain an impression whether this effect is big enough to override potential negative “side effects”.

## METHODS

In this retrospective cohort study, we compared cumulative SARS-CoV-2 infection rates in sports classes (with limited use of face masks) and non-sports classes (with the consequent use of face masks). The study was registered in the German Clinical Trial Register (ID DRKS00029061) and approved by the Research Ethics Committee at the University of Graz, Styria, Austria (GZ. 39/70/63 ex 2021/22).

### Selection of school and participants

A school campus in Klagenfurt, Austria was selected, consisting of a secondary school with a focus on “development for competitive sports”, and a parallel general school branch (GB). Children/adolescents attending the branch with a sports focus (SF) were allowed to participate in sports and other physical activity without restriction, due to the guidelines and safety measures for competitive sports in Austria.^19^ In SF classes, students completed a two-hour sports lesson three times weekly, mostly indoors.

Students attending the GB branch were allowed to carry out physical activities and sports only under strict restrictions in compliance with existing COVID-19 mitigation measures.^19^ At the onset of data collection, 616 students (421 in GB and 195 in SF) were attending secondary school education at the campus. They, respectively their parents were asked to provide information about SARS-CoV-2 infection during the period of interest. A total of 213 legal guardians (for children < 14 years) and 401 students (≥ 14 years) gave written consent to participate in the study. Two students (0.3%) did not want to participate in the study, both of them from the GB classes.

### Definition of different periods based on dominant virus variants and mitigation measures

In Austria, a detailed school safety concept was mandated by the Federal Ministry of Education, Science, and Research in August 2021 for the 2021/22 school year in order to ensure largely unrestricted school operations during the COVID-19 pandemic. Three safety levels with different mitigation measures were defined to be adapted to the regional SARS-CoV-2 infection situation.^19^

In Sept and Oct 2021, very low 7-day incidence of SARS-CoV-2 infections occurred in the region around the school campus. Therefore, after a 3-week security phase (with one polymerase chain reaction (PCR) test and two rapid antigen tests weekly), security level 1 was enacted (students had the option of voluntary testing at school via a rapid antigen test). From Nov 2, 2021 to Feb 28, 2022, security level 3 was enacted, meaning that students were required to repeatedly perform a SARS-CoV-2 screening test three times a week (two polymerase chain reaction (PCR) tests and one rapid antigen test), to keep a safety distance of 1 meter, and to wear an FFP-2 mask throughout the school building. If infection with SARS-CoV-2 was detected by PCR testing, students had to stay at home for 10 days. Subsequently, students who tested positive were excluded from further testing for the following 90 days.^20,21^ From Feb 28 onwards, the strict mitigation measures were relaxed and students were no longer requested to wear FFP-2 masks in classrooms. However, PCR and antigen testing were continued (two polymerase chain reaction (PCR) tests and one rapid antigen test).^22^ During the study period, two variants of SARS-CoV-2 circulated in Austria that were classified as variants of concern by the World Health Organization (WHO).^23^ The delta variant B.1.617.2 was predominant until December 2021^24,25^, while from January 2022 onwards the Omicron variant BA.1^26^ was the dominant variant.^24,25^

Based on the different mitigation measures imposed and the dominant variants in the various periods, the study period was divided into four periods of relatively equal length (see also eTable 1):

P1 (September 13, 2021 to October 31, 2021): the predominant SARS-CoV-2 variant = delta (B.1.617.2); the low 7-day incidence of SARS-CoV-2 infections, security phase, and security level 1;

P2 (November 1, 2021 to December 31, 2021): the predominant SARS-CoV-2 variant = delta (B.1.617.2); a high 7-day incidence of SARS-CoV-2 infections, security level 3;

P3 (January 1, 2022 to February 28, 2022): the predominant SARS-CoV-2 variant = Omicron (BA.1); a very high 7-day incidence of SARS-CoV-2 infections, security level 3;

P4 (March 1, 2022 to April 30, 2022): the predominant SARS-CoV-2 variant = Omicron (BA.2/BA.3); a very high 7-day incidence of SARS-CoV-2 infections, and relaxed security level 3 without wearing FFP2 masks in classes.

### Procedures

In May and June 2022, the students provided information on whether and when they had had an infection with SARS-CoV-2 detected by PCR test between September 13, 2021 and April 30, 2022.

Dichotomous data (the detection of SARS-CoV-2 by PCR testing: yes or no) were generated for each day of the study period for all participants. In a second step, potential secondary cases (infection) in the class were identified to estimate the secondary attack rate considering the current knowledge about mean generation time (GT).

Mean GT is the time interval between a reported infection of a primary case (infector) and potential secondary cases (infected).^27^ Previous studies have reported different GTs for different variants of SARS-CoV-2.^28-31^ Zhang et al.^28^ reported a mean GT of 2.9 days for B.1.617.2, while Hart et al.^29^ reported a mean intrinsic GT of 4.7 days and a mean household GT of 3.2 days for this variant. Ito et al.^30^ reported a reduction in mean GT for the Omicron variant BA.1 compared with the delta variant B.1.617.2 (GT for BA.1 = 0.44 to 0.46 times delta), and similar results were reported by Manica et al.^31^.

Based on these reports and the fact that GTs for potential secondary cases (infections) are difficult to measure^27^, different intervals (2, 4, 6, or 8 days) for mean GT were used in our analyses. For all reported SARS-CoV-2 infections, potential secondary cases (infections) in the same classroom were identified for each assumed GT (GT = 2, 4, 6, or 8 days) and dichotomous data (yes or no) were generated for each day and student.

### Outcomes

The primary outcomes were SARS-CoV-2 infection dynamics (a cumulative percentage of students with a SARS-CoV-2 infection), which were compared between the SF and GB school classes. Using reported SARS-Cov-2 infections, daily 7-day class incidence was calculated by dividing the number of positive test results in the previous 7 days by the number of students attending class and extrapolating the obtained 7-day class density to 100,000 individuals.

As mentioned, potential secondary cases in school classes were identified using different GTs (2, 4, 6, or 8 days), and odds ratios were calculated for students in SF and GB classes. Secondary analyses were performed for subgroups based on school grade (middle school (M.S.): age 10-16 years (mean 13.1 [95%CI=12.9-13.2 years]); 4-year high school (H.S.): age 14-20 years (mean 16.9 [95%CI=16.8-17.1 years])) and sex (boys; girls).

### Statistical analysis

Descriptive statistics were calculated; continuous variables are expressed as mean (M) and standard deviation (SD), and categorial variables as absolute values (n) and percentages (%), and no data imputation was performed.

We used a chi-square test (X^2^) or a Fisher’s exact test where appropriate to test for differences in cumulative percentages of students with a SARS-CoV-2 infection in SF and GB classes. Phi coefficients (φ) were calculated to estimate the correlation strength between stringent mitigation measures in the SF and GB school classes.

Additionally, a binary logistic regression analysis was performed to assess the relationship between class membership (SF or GB) and the cumulative percentage of students with a SARS-CoV-2 infection (CPI) or the potential SARS-CoV-2 cases (infections) in classroom settings. Coefficients obtained from binary logistic regression were expressed as odds ratios (OR) with 95% confidence intervals. Due to a lack of variance homogeneity, we used the Welch test to analyze differences in 7-day incidences.

All tests were two-sided, with a p-value of <0.05 considered statistically significant. Phi (φ) according to Cohen^32^ was used to determine the effect size (≥0.1, small; ≥0.3, medium; and ≥0.5, large). All statistical calculations were performed using SPSS Version 27 (IBM Corp. Released 2020. IBM SPSS Statistics for Windows, Armonk, NY: IBM Corp).

## RESULTS

Among the 614 students (age: 15.1 ± 2.3 years, 43.2% female) included in this analysis, 195 students (31.8%) attended school classes with a sports focus (SF) and were allowed to participate in sports without restrictions during the study period. Significantly fewer girls were in SF classes than in GB classes (SF: ♀=25.1%, GB: ♀=51.6%; p≤.001). Students in SF classes were slightly younger than those in GB classes (SF: 14.8 years; GB: 15.3 years; p = 0.006) (eTable 2).

In total, 76 students (12.4%) had been infected with SARS-CoV-2 before the study period (before September 13, 2021). More previous infections were reported by older students (H.S.) in SF classes than by students in parallel GB classes (H.S. = SF: 22.9%; GB: 11.8%; and p=0.011). No differences in previous infections were reported by younger students (M.S. = SF: 10.1%; GB: 8.9%; p=0.75).

### Infection status—changes over time

#### CPI— Cumulative percentage of students with a SARS-CoV-2 infection

No significant differences were found in the CPI by the end of the time period 1 (P1) between SF and GB classes, and overall, the CPI levels were very low (SF: 2.6%; GB 1.2%; p = 0.30; and φ=0.05) (Table 1, Figure 1).

**Table 1:** Cumulative percentage of students with a SARS-CoV-2 infection: students in classes with GB vs students in SF classes

| Variable | Time period | All (n=614) |  | p Value | φ | OR (95% CI) |
| --- | --- | --- | --- | --- | --- | --- |
|  |  | GB (n=419) | SF (n=185) |  |  |  |
| CU% SARS-CoV-2 ..., No. [%] | ...Sep.21 (P1) | 1 (0.2%) | 2 (1.0%) | .24 | .05 | 4.33 (0.39 to 48.06) |
|  | ...Oct.21 (P1) | 5 (1.2%) | 5 (2.6%) | .30 | .05 | 2.17 (0.62 to 7.62) |
|  | ...Nov.21 (P2) | 26 (6.2%) | 29 (14.9%) | <.001 | .14 | 2.64 (1.51 to 4.62) |
|  | ...Dec.21 (P2) | 38 (9.1%) | 32 (16.4%) | .008 | .11 | 1.97 (1.19 to 3.26) |
|  | ...Jan.22 (P3) | 81 (19.3%) | 85 (43.6%) | <.001 | .25 | 3.22 (2.22 to 4.68) |
|  | ...Feb.22 (P3) | 149 (35.6%) | 115 (59.0%) | <.001 | .22 | 2.61 (1.84 to 3.69) |
|  | ...Mar.22 (P4) | 215 (51.3%) | 120 (61.5%) | .018 | .10 | 1.52 (1.07 to 2.15) |
|  | ...Apr.22 (P4) | 229 (54.7%) | 124 (63.6%) | .037 | .09 | 1.45 (1.02 to 2.06) |
|  |  | M.S. (n=289) |  |  |  |  |
|  |  | GB (n=190) | SF (n=99) |  |  |  |
| CU% SARS-CoV-2 ..., No. [%] | ...Sep.21 (P1) | 0 (0.0%) | 1 (1.0%) | .34 | .08 | INC |
|  | ...Oct.21 (P1) | 2 (1.1%) | 2 (2.0%) | .61 | .04 | 1.94 (0.27 to 13.97) |
|  | ...Nov.21 (P2) | 9 (4.7%) | 15 (15.2%) | .002 | .18 | 3.59 (1.51 to 8.54) |
|  | ...Dec.21 (P2) | 15 (7.9%) | 18 (18.2%) | .009 | .15 | 2.59 (1.24 to 5.40) |
|  | ...Jan.22 (P3) | 33 (17.4%) | 40 (40.4%) | <.001 | .25 | 3.23 (1.86 to 5.59) |
|  | ...Feb.22 (P3) | 70 (36.8%) | 60 (60.6%) | <.001 | .23 | 2.64 (1.60 to 4.35) |
|  | ...Mar.22 (P4) | 101 (53.2%) | 64 (64.6%) | .06 | .11 | 1.61 (0.98 to 2.66) |
|  | ...Apr.22 (P4) | 107 (56.3%) | 66 (66.7%) | .09 | .10 | 1.55 (0.94 to 2.58) |
|  |  | H.S. (n=325) |  |  |  |  |
|  |  | GB (n=229) | SF (n=96) |  |  |  |
| CU% SARS-CoV-2 ..., No. [%] | ...Sep.21 (P1) | 1 (0.4%) | 1 (1.0%) | .50 | .04 | 2.40 (0.15 to 38.77) |
|  | ...Oct.21 (P1) | 3 (1.3%) | 3 (3.1%) | .37 | .06 | 2.43 (0.48 to 12.26) |
|  | ...Nov.21 (P2) | 17 (7.4%) | 14 (14.6%) | .045 | .11 | 2.13 (1.00 to 4.52) |
|  | ...Dec.21 (P2) | 23 (10.0%) | 14 (14.6%) | .24 | .07 | 1.53 (0.75 to 3.12) |
|  | ...Jan.22 (P3) | 48 (21.0%) | 45 (46.9%) | <.001 | .26 | 3.32 (1.99 to 5.55) |
|  | ...Feb.22 (P3) | 79 (34.5%) | 55 (57.3%) | <.001 | .21 | 2.55 (1.56 to 4.15) |
|  | ...Mar.22 (P4) | 114 (49.8%) | 56 (58.3%) | .16 | .08 | 1.41 (0.87 to 2.29) |
|  | ...Apr.22 (P4) | 122 (53.3%) | 58 (60.4%) | .24 | .07 | 1.34 (0.83 to 2.17) |

Data are No. (%), M.S. = middle school (students aged $13.1 \pm 1.3$ years old), H.S. = 4-year high school (students aged $16.9 \pm 1.2$ years old), GB = students in school classes with a general school branch, SF = students in school classes with sport focus, φ = Effect size Phi, OR = odds ratio (reference group: students without sport focus), CI = confidence interval, CU% SARS-CoV-2 = cumulative percentage of students with a SARS-COV-2 infections at the End of..., SARS-CoV-2 = severe acute respiratory syndrome coronavirus 2, P1 = period 1 (September 13, 2021 to October 31, 2021) P2 = period 2 (November 1, 2021 to December 31, 2021), P3 = period 3 (January 1, 2022 to February 28, 2022), P4 = period 4 (March 1, 2022 to April 30, 2022), INC = insufficient number of potential infection cases.

**Figure 1.**
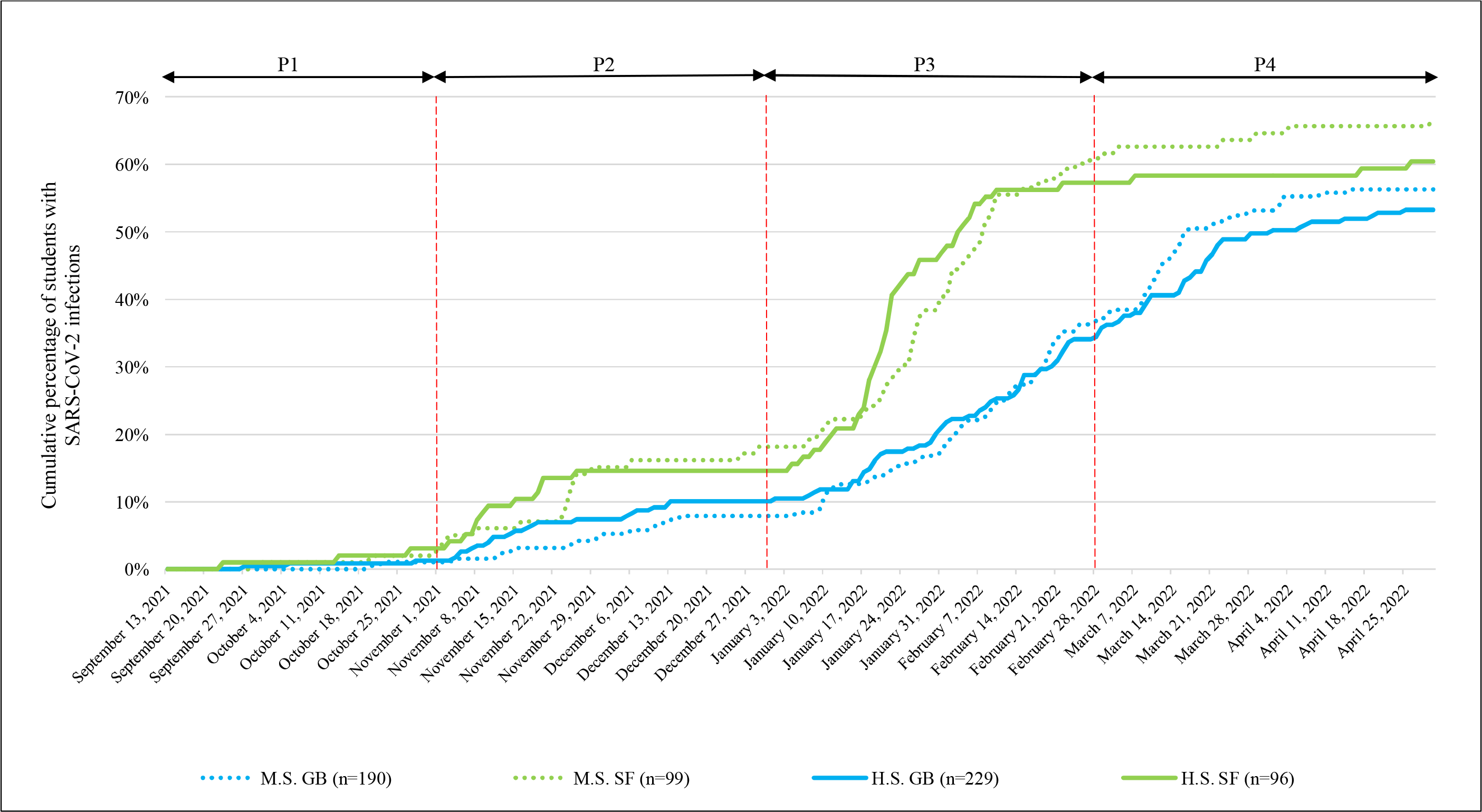
Cumulative percentage of students with SARS-CoV-2 infections SARS-CoV-2 = severe acute respiratory syndrome coronavirus 2, P1 = period 1 (September 13, 2021 to October 31, 2021) P2 = period 2 (November 1, 2021 to December 31, 2021), P3 = period 3 (January 1, 2022 to February 28, 2022), P4 = period 4 (March 1, 2022 to April 30, 2022), M.S. = middle school (Children aged 13.1 ± 1.3 years old), H.S. = 4-year high school (Children aged 16.9 ± 1.2 years old), GB = students in school classes with a general school branch, SF = students in school classes with sport focus.

In time period 2 (P2), the CPI increased and moderate differences were observed (end of Dec 2021: SF: 16.4%; GB: 9.1%; p = 0.008, φ=0.11). For students in SF school classes, the odds of becoming infected with SARS-CoV-2 increased during P2 (the end of Nov 2021: OR = 2.64 (95% CI, 1.51-4.62); the end of Dec 2021: OR = 1.97 (95% CI, 1.19-3.26)). ORs were higher for younger (M.S.) students (the end of Nov 2021: OR = 3.59 (95% CI, 1.51-8.54); the end of Dec 2021: OR = 2.59 (95% CI, 1.24-5.40)) than older (H.S.) students (the end of Nov 2021: OR = 2.13 (95% CI, 1.00-4.52); the end of Dec 2021: OR = 1.53 (95% CI, 0.75-3.12)) (Table 1, Figure 1).

CPI increased dramatically in time period 3 (P3) due to the Omicron wave. By the end of February 2022, greatly increased CPI in both branches went along with a significant difference between the school focuses (SF: 60.6%; GB: 36.8%; p <.001; and φ=0.23). At the end of February 2022, students in the SF school classes exhibited a 2.6-fold increased probability of having been infected with SARS-CoV-2 (end of Feb 2022: OR = 2.61 (95% CI, 1.84-3.69)) (Table 1, Figure 1).

By the end of March 2022, this difference decreased, with students in GB school classes showing significantly higher CPI compared to the end of the previous month (SF: 61.5%; GB: 53.2%; p=0.018, φ=0.10). This trend continued until the end of April 2022 (SF: 63.1%; GB: 54.7%; p=0.037, φ=0.09) and could also be observed in different school grades (M.S. and H.S.) (Table 1, Figure 1). Detailed information on different CPI trends in subgroups are reported in the supplements (eTable 3-7, eFigure 1).

#### Mean 7-day incidence for infection with SARS-CoV-2 (M7D-I SARS-CoV-2)

M7D-I SARS-CoV-2 showed significant differences (p<.001) between school focuses in all four time periods. In P1, P2, and P3, higher (p<.001) M7D-I SARS-CoV-2 was observed in SF school classes. During P4, the situation reversed, and significantly higher (p<.001) M7D-I SARS-CoV-2 were detected in GB school classes (Table 2, Figure 2). Detailed information on trends in M7D-I SARS-CoV-2 are reported in the Supplementary Materials (eTable 8-11).

**Table 2.** Mean 7-day incidences in classrooms for SARS-CoV-2 infections per 100,000 inhabitants: students in GB classes vs students in classes with SF

| Variable | Time period | Mean AGES 7-day I (5 to 14) | M.S. (n=289) |  | t<sup>a</sup> | P Value | p-lvl | M.D. | 95% CI |  |
| --- | --- | --- | --- | --- | --- | --- | --- | --- | --- | --- |
|  |  |  | GB (n=190) | SF (n=99) |  |  |  |  | Lo | Up |
| Mean 7-day I (CR) | P1 | 370.02 | 118.85 ± 202.19 | 251.62 ± 251.08 | -4.549 | <.001 | *** | -132.77 | -190.40 | -75.15 |
|  | P2 | 1521.82 | 783.81 ± 732.41 | 1903.99 ± 564.43 | -14.412 | <.001 | *** | -1120.17 | -1273.26 | -967.08 |
|  | P3 | 2797.19 | 3593.86 ± 1502.88 | 5319.90 ± 653.95 | -13.558 | <.001 | *** | -1726.04 | -1976.64 | -1475.44 |
|  | P4 | 2361.77 | 2372.95 ± 1155.04 | 906.38 ± 870.70 | 12.105 | <.001 | *** | 1466.57 | 1227.96 | 1705.19 |
|  |  | Mean AGES 7-day I (15 to 24) | H.S. (n=325) |  |  |  |  |  |  |  |
|  |  |  | GB (n=229) | SF (n=96) |  |  |  |  |  |  |
| Mean 7-day I (CR) | P1 | 268.88 | 196.43 ± 335.04 | 404.16 ± 507.97 | -3.685 | <.001 | *** | -207.73 | -319.25 | -96.21 |
|  | P2 | 962.70 | 1030.43 ± 768.68 | 1370.41 ± 1145.24 | -2.668 | <.001 | *** | -339.98 | -592.08 | -87.88 |
|  | P3 | 2497.02 | 3141.79 ± 994.91 | 5324.14 ± 1282.66 | -14.897 | <.001 | *** | -2182.35 | -2471.88 | -1892.82 |
|  | P4 | 2185.20 | 2488.14 ± 1203.69 | 602.6 ± 582.11 | 18.992 | <.001 | *** | 1885.55 | 1690.21 | 2080.88 |

a = Due to lack of variance homogeneity, we used the Welch test.
Data are mean (SD), SD = standard deviation; AGES = Austrian Agency for Health and Food Safety, 7-day I (5 to 14) = 7 days incidence for SARS-CoV-2 infections per 100,000 inhabitants in Carinthia (AUT) for children aged 5 to 14 years (https://covid19-dashboard.ages.at/), 7-day I (15 to 24) = 7 days incidence for SARS-CoV-2 infections per 100,000 inhabitants in Carinthia (AUT) for adolescent aged 15 to 24 years (https://covid19-dashboard.ages.at/), AUT = Austria, M.S. = middle school (Students aged 13.1 ± 1.3 years old), H.S. = 4-year high school (Students aged 16.9 ± 1.2 years old), GB = students in school classes with a general school branch, SF = students in school classes with sport focus, t = test statistic welch test; p-lvl (P Value level) \* = P <.05, \*\* = P <.01, \*\*\* = P <.001, M.D. = Mean Difference, CI = Confidence Interval of the Difference, Lo = Lower, Up = Upper, I(CR) = incidence in classroom, CR = classroom, P1 = period 1 (13. September to 31. October 2021), P2 = period 2 (1. November to 31. December 2021), P3 = period 3 (1. January to 28. February 2022), P4 = period 4 (1. March and 30. April 2022).

**Figure 2.**
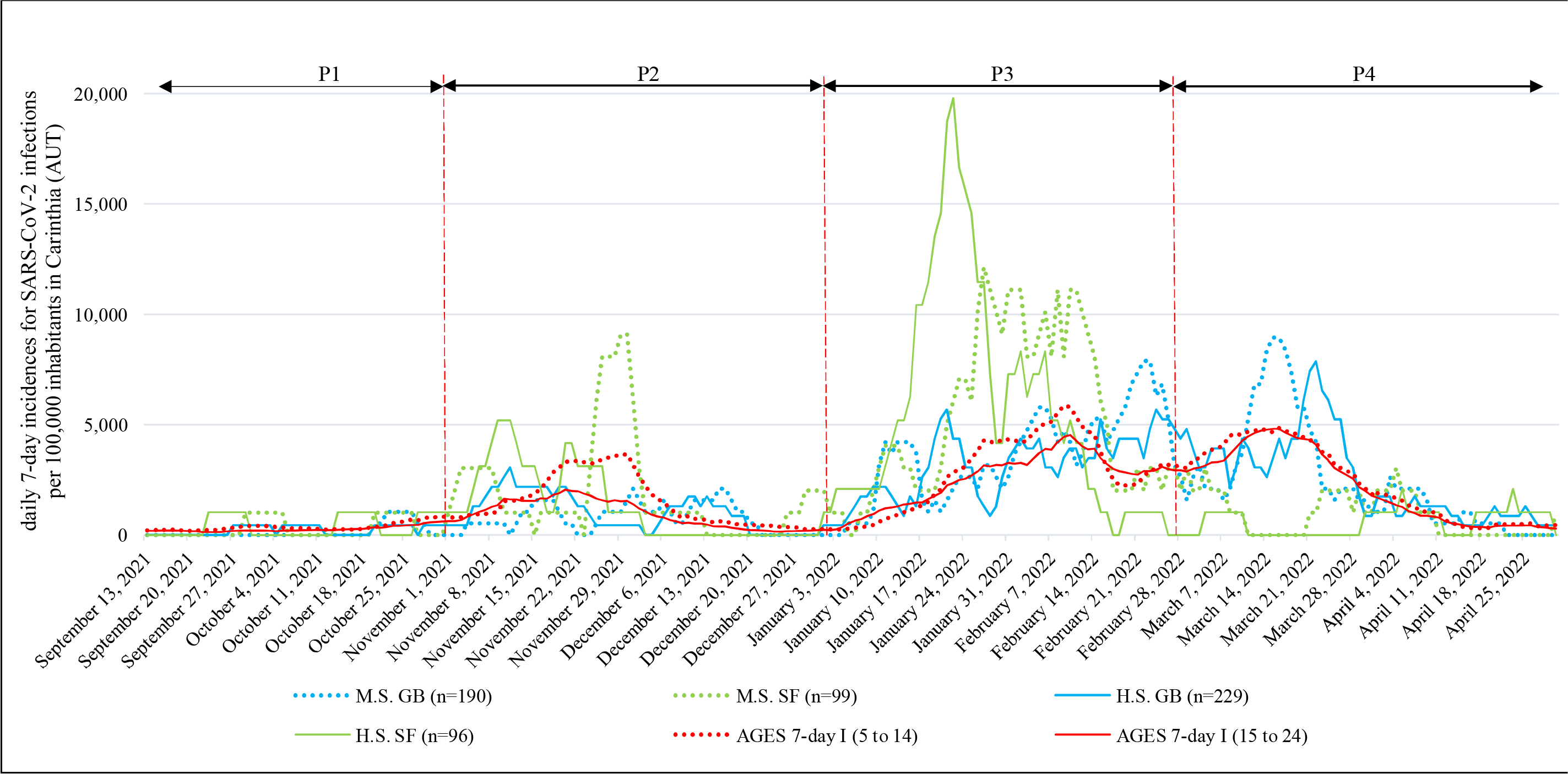
Differences in 7-day incidence of SARS-CoV-2 infections per 100,000 inhabitants in Carinthia (AUT) and among students of secondary school age with or without a sport focus. SARS-CoV-2 = severe acute respiratory syndrome coronavirus 2, P1 = period 1 (September 13, 2021 to October 31, 2021) P2 = period 2 (November 1, 2021 to December 31, 2021), P3 = period 3 (January 1, 2022 to February 28, 2022), P4 = period 4 (March 1, 2022 to April 30, 2022), M.S. = middle school (Children aged 13.1 ± 1.3 years old), H.S. = 4-year high school (Children aged 16.9 ± 1.2 years old), GB = students in school classes with a general school branch, SF = students in school classes with sport focus, AGES = Austrian Agency for Health and Food Safety (https://covid19-dashboard.ages.at/), 7-day I (5 to 14) = 7 days incidence for SARS-CoV-2 infections per 100,000 inhabitants in Carinthia (AUT) for students aged 5 to 14 years, 7-day I (15 to 24) = 7 days incidence for SARS-CoV-2 infections per 100,000 inhabitants in Carinthia (AUT) for people aged 15 to 24 years, AUT = Austria.

#### Potential SARS-CoV-2 infection in school classes (PI-SARS-CoV-2)

When PI-SARS-CoV-2 was analyzed, small effects were found in P2, P3, and P4 regardless of which hypothesized GT (mean generation time) had been used. Due to the limited number of participants, GT for 2 days during P1, P2, and P4 partially identified insufficient numbers of potential infection cases. Regardless of which GT was used, there were no significant changes in the overall conclusion of which school branch was more likely to be infected with SARS-CoV-2 (Table 3). In P2 and P3, the odds of PI-SARS-CoV-2 were higher in SF classes (hypothesized GT for 4 days= P2: OR = 2.78 (95% CI, 1.08-7.15); P3: OR = 4.03 (95% CI, 2.56-6.31)). These changes in P4, where the odds of PI- SARS-CoV-2 were significantly reduced in SF classes, were in contrast to those in P2 and P3 (hypothesized GT for 4 days = P4: OR = 0.05 (95% CI, 0.01-0.39)) (Table 3). Additional results for differentially hypothesized GTs are reported in the supplements (eTable 12-17).

**Table 3:**
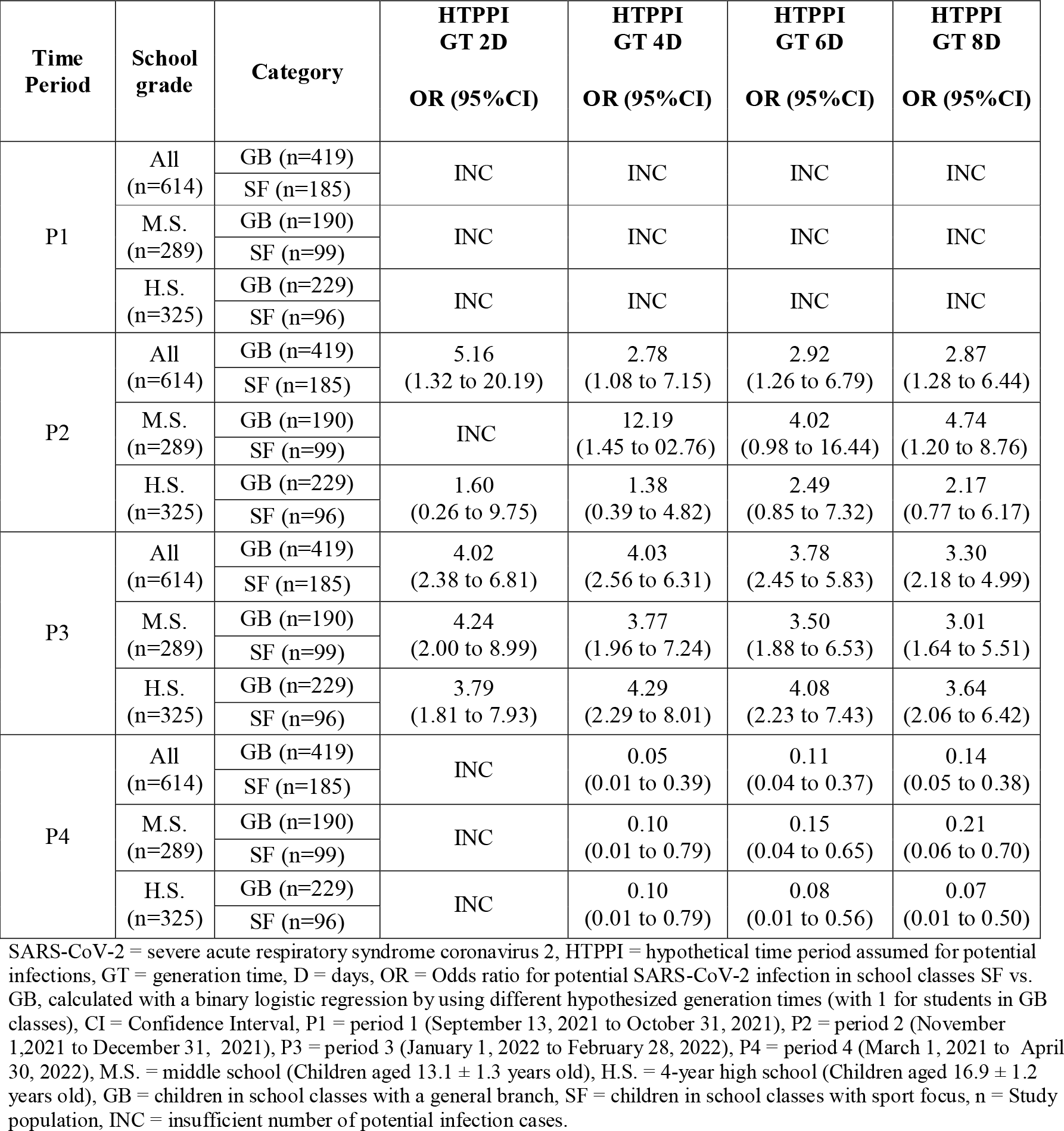
Potential SARS-CoV-2 infection in school classes: SF vs. GB, using different hypothesized generation time.

| Time Period | School grade | Category | HTPPI GT 2D<br>OR (95%CI) | HTPPI GT 4D<br>OR (95%CI) | HTPPI GT 6D<br>OR (95%CI) | HTPPI GT 8D<br>OR (95%CI) |
| --- | --- | --- | --- | --- | --- | --- |
| P1 | All (n=614) | GB (n=419) | INC | INC | INC | INC |
|  |  | SF (n=185) |  |  |  |  |
|  | M.S. (n=289) | GB (n=190) | INC | INC | INC | INC |
|  |  | SF (n=99) |  |  |  |  |
|  | H.S. (n=325) | GB (n=229) | INC | INC | INC | INC |
|  |  | SF (n=96) |  |  |  |  |
| P2 | All (n=614) | GB (n=419) | 5.16<br>(1.32 to 20.19) | 2.78<br>(1.08 to 7.15) | 2.92<br>(1.26 to 6.79) | 2.87<br>(1.28 to 6.44) |
|  |  | SF (n=185) |  |  |  |  |
|  | M.S. (n=289) | GB (n=190) | INC | 12.19<br>(1.45 to 02.76) | 4.02<br>(0.98 to 16.44) | 4.74<br>(1.20 to 8.76) |
|  |  | SF (n=99) |  |  |  |  |
|  | H.S. (n=325) | GB (n=229) | 1.60<br>(0.26 to 9.75) | 1.38<br>(0.39 to 4.82) | 2.49<br>(0.85 to 7.32) | 2.17<br>(0.77 to 6.17) |
|  |  | SF (n=96) |  |  |  |  |
| P3 | All (n=614) | GB (n=419) | 4.02<br>(2.38 to 6.81) | 4.03<br>(2.56 to 6.31) | 3.78<br>(2.45 to 5.83) | 3.30<br>(2.18 to 4.99) |
|  |  | SF (n=185) |  |  |  |  |
|  | M.S. (n=289) | GB (n=190) | 4.24<br>(2.00 to 8.99) | 3.77<br>(1.96 to 7.24) | 3.50<br>(1.88 to 6.53) | 3.01<br>(1.64 to 5.51) |
|  |  | SF (n=99) |  |  |  |  |
|  | H.S. (n=325) | GB (n=229) | 3.79<br>(1.81 to 7.93) | 4.29<br>(2.29 to 8.01) | 4.08<br>(2.23 to 7.43) | 3.64<br>(2.06 to 6.42) |
|  |  | SF (n=96) |  |  |  |  |
| P4 | All (n=614) | GB (n=419) | INC | 0.05<br>(0.01 to 0.39) | 0.11<br>(0.04 to 0.37) | 0.14<br>(0.05 to 0.38) |
|  |  | SF (n=185) |  |  |  |  |
|  | M.S. (n=289) | GB (n=190) | INC | 0.10<br>(0.01 to 0.79) | 0.15<br>(0.04 to 0.65) | 0.21<br>(0.06 to 0.70) |
|  |  | SF (n=99) |  |  |  |  |
|  | H.S. (n=325) | GB (n=229) | INC | 0.10<br>(0.01 to 0.79) | 0.08<br>(0.01 to 0.56) | 0.07<br>(0.01 to 0.50) |
|  |  | SF (n=96) |  |  |  |  |

SARS-CoV-2 = severe acute respiratory syndrome coronavirus 2, HTPPI = hypothetical time period assumed for potential infections, GT = generation time, D = days, OR = Odds ratio for potential SARS-CoV-2 infection in school classes SF vs. GB, calculated with a binary logistic regression by using different hypothesized generation times (with 1 for students in GB classes), CI = Confidence Interval, P1 = period 1 (September 13, 2021 to October 31, 2021), P2 = period 2 (November 1, 2021 to December 31, 2021), P3 = period 3 (January 1, 2022 to February 28, 2022), P4 = period 4 (March 1, 2021 to April 30, 2022), M.S. = middle school (Children aged $13.1 \pm 1.3$ years old), H.S. = 4-year high school (Children aged $16.9 \pm 1.2$ years old), GB = children in school classes with a general branch, SF = children in school classes with sport focus, n = Study population, INC = insufficient number of potential infection cases.

## DISCUSSION

To our knowledge, our study is the first to assess the impact of wearing face masks in classroom settings. Our results show that under otherwise strict mitigation measures, students allowed to participate in sports without restriction (SF group) were at a significantly higher risk of being infected by SARS-CoV-2 than those following general strict mitigation measures at school. This is in line with the results of studies reporting the clear efficacy of wearing FFP2 or any other face masks, with both leading to a reduction in viral (variant) spreading.^33,34^

To the best of our knowledge, the long-term effects of continued mask wearing have been investigated in only a few studies to date.^35-38^ These suggest that the continued wearing of face masks may also have negative consequences, and these must be carefully balanced against the advantage of reduced infection rates. In this context, the question arises of whether the consequent routine use of face masks is able to reduce long-term infection rates or merely postpones numerous infections. The latter could reduce the usefulness of long-term mask obligations, together with the reduced pathogenicity of the virus variant that is circulating. In late February 2022, an increasing number of studies reported that Omicron variants led to less severe disease, and reduced hospitalization rates. In response, stringent mitigation measures were relaxed worldwide.^39,40^ Our data show that this (at least in school settings) apparently led to “catch-up” infections in those thus far protected from infection with SARS-CoV-2. By the end of our data collection (April 2022), only a small difference in cumulative infection rates was observed between students with strict mask wearing rules during the high-incidence period and those without.

This leads to the question when, for whom, and which face masks should be made obligatory in school settings, and especially for how long. These questions must be answered by medical research and should no longer be a matter of individual/local/regional decision-making. Although no major physical/physiological negative side effects of face mask use have been proven for healthy infants so far^36^, continued or even long-term use may have a negative impact on long-term psychosocial health in growing and developing individuals^13-15,41^. In an attempt to avoid viral transmission whenever and wherever possible, this aspect was seemingly underrepresented in systematic COVID-19 research. Our study demonstrates that wearing face masks is effective at reducing viral transmission. This benefit is, however, limited in the long run. Thus, the benefit of viral transmission reduction must be carefully balanced against potential negative effects on mental and social health.

### Strengths and limitations

Due to the limited number of school classes without strict mitigation measures during the school year 2021/2022, no random representative sample selection was possible. However, the sample size was large and the participation rate was high (99.7%).

Another limitation of our study is that the actual chain of SARS-CoV-2 transmissions cannot be detected flawlessly in practice. In fact, transmission could also have taken place outside school by personal contacts during leisure time. This limitation, however, holds true for almost all studies dealing with secondary attack estimations.

### Implications

Regardless of the limitations, our results clearly show the “catch-up” of infections in response to the relaxation of strict mitigation measures. Thus, the temporary obligation of face mask use in schools can postpone a number of secondary infections but has a very limited effect on the long-term avoidance of these infections. This finding should be considered regarding further decisions concerning obligatory face mask use in school settings.

## Conclusion

Our study shows that a number of infections with SARS-CoV-2 are delayed, but they cannot be prevented in the long run by wearing face masks. Therefore, the obligatory use of face masks in schools may be understood as an epidemiological measure to flatten SARS-CoV-2 peaks rather than to protect individuals. Since healthy school children are rarely severely affected by COVID on the one hand, but may experience negative psychosocial consequences on the other hand by continued face mask use, the advantage of (temporarily) reduced virus transmission must be carefully balanced against the potential negative consequences on psychosocial development and mental health.

## Supporting information

supplements Jarnig et al

## Data Availability

All data produced in the present work are contained in the manuscript

## Author Contributions

All authors had full access to all the data in the study and take responsibility for the integrity of the data and the accuracy of the data analysis.

**Concept and Design:** Jarnig, Kerbl, and van Poppel.

**Acquisition, Analysis, or Interpretation of Data:** Jarnig, Kerbl, and van Poppel.

**Drafting of the Manuscript:** Jarnig.

**Critical Revision of the Manuscript for Important Intellectual Content**: Jarnig, Kerbl, and van Poppel.

**Statistical Analysis:** Jarnig and van Poppel.

**Obtained Funding:** Jarnig.

**Administrative, Technical, or Material Support:** Jarnig.

**Supervision:** Jarnig, Kerbl, and van Poppel.

## Conflict of Interest Disclosures

We declare no competing interests.

## Funding/Support

The Austrian Federal Ministry of Education, Science, and Research (GZ: 2022-0.333.520).

## Role of the funding source

The funder of the study had no role in the study design, data collection, data analysis, data interpretation, or writing of the report. All authors had full access to the data in the study and had final responsibility for the decision to submit the study for publication.

## Data Sharing Statement

Data collected for the study, including individual participant data, will not be made available to others since subsequent follow-up investigations are in progress. When all follow-up investigations are finished, data might be made available upon reasonable request.

## Additional Contributions

This study (and GJ) was funded by the Austrian Federal Ministry of Education, Science, and Research (GZ: 2022-0.333.520) and organized by the scientific freelancer GJ. We would like to thank all participants and their legal guardians for providing the data.

## Notes

### Competing Interest Statement

The authors have declared no competing interest.

### Funding Statement

This study was funded by the Austrian Federal Ministry of Education, Science, and Research
(GZ: 2022-0.333.520).

### Author Declarations

The study was registered in the German Clinical Trial Register (ID DRKS00029061) and approved by the Research Ethics Committee at the University of Graz, Styria, Austria (GZ. 39/70/63 ex 2021/22).

### Summary of Updates

In the abstract, a typographical error was corrected and a word repetition was corrected.

