## supplements Jarnig et al for "Effects of wearing FFP2 masks on SARS-CoV-2 infection rates in classrooms"

**eTable 1.** Description of the 4 time periods within the course of the study.

**eTable 2.** Sample characteristics for total study population and subgroups middle and high school

**Table 3:** Cumulative percentage of students with a SARS-CoV-2 infection: students in classes with GB vs students in SF classes for subgroups sex.

**eTable 4.** Detailed increase of cumulative numbers and percentage of students with a SARS-CoV-2 infection for P1 (Sep.21 and Oct.21).

**eTable 5.** Detailed increase of cumulative numbers and percentage of students with a SARS-CoV-2 infection for P2 (Nov.21 and Dec.21).

**eTable 6.** Detailed increase of cumulative numbers and percentage of students with a SARS-CoV-2 infection for P3 (Jan.22 and Feb.22).

**eTable 7.** Detailed increase of cumulative numbers and percentage of students with a SARS-CoV-2 infection for P4 (Mar.22 and Apr.22).

**eTable 8.** Detailed 7-day incidences for detected SARS-CoV-2 infections for P1 (Sep.21 and Oct.21).

**eTable 9.** Detailed 7-day incidences for detected SARS-CoV-2 infections for P2 (Nov.21 and Dec.21).

**eTable 10.** Detailed 7-day incidences for detected SARS-CoV-2 infections for P3 (Jan.22 and Feb.22).

**eTable 11.** Detailed 7-day incidences for detected SARS-CoV-2 infections for P4 (Mar.22 and Apr.22).

**eTable 12:** Potential SARS-CoV-2 infection in school classes GB vs. SF classes, using hypothesized generation time - 2 days.

**eTable 13:** Potential SARS-CoV-2 infection in school classes GB vs. SF classes, using hypothesized generation time - 4 days.

**eTable 14:** Potential SARS-CoV-2 infection in school classes GB vs. SF classes, using hypothesized generation time - 6 days.

**eTable 15:** Potential SARS-CoV-2 infection in school classes GB vs. SF classes, using hypothesized generation time - 8 days.

**eTable 16.** Binary logistic regression for Potential SARS-Cov-2 contagions in the classroom and generation times 4 and 6, for subgroup sex .

**eTable 17.** Binary logistic regression for Potential SARS-Cov-2 contagions in the classroom and generation times 2 and 8, for subgroup sex .

**eFigure 1.** Cumulative percentage of children with SARS-CoV-2 infections, for subgroup sex.

This supplementary material has been provided by the authors to give readers additional information about their work.

**eTable 1.** Description of the 4 time periods within the course of the study.

| Time period | date range | Mitigation measures at school not included in physical education classes | | School sports | | Predominant SARS-CoV-2 variant | AGES | |
|---|---|---|---|---|---|---|---|---|
| | | GB | SF | GB | SF | | Mean 7-day I (5 to 14) | Mean 7-day I (15 to 24) |
| P1 | September 13, 2021 to October 31, | A, B | A, B | Outdoor without restrictions, indoor with restrictions | No restrictions | B.1.617.2 | 370.02 | 268.88 |
| P2 | November 1, 2021 to December 31, | C | C | Outdoor with restrictions, indoor with strict restrictions and wearing FFP2 masks | No restrictions | B.1.617.2 | 1521.82 | 962.70 |
| P3 | January 1, 2022 to February 28, 2022 | C | C | Outdoor with restrictions, indoor with strict restrictions and wearing FFP2 masks | No restrictions | BA.1 | 2797.19 | 2497.02 |
| P4 | March 1, 2022 to April 30, 2022 | D | D | Outdoor without restrictions, indoor with restrictions | No restrictions | BA.2/BA.3 | 2361.77 | 2185.20 |

SARS-CoV-2 = severe acute respiratory syndrome coronavirus 2, GB = students in school classes with a general school branch, SF = students in school classes with sport focus, AGES = Austrian Agency for Health and Food Safety, 7-day I (5 to 14) = 7 days incidence for SARS-CoV-2 infections per 100,000 inhabitants in Carinthia (AUT) for children aged 5 to 14 years (https://covid19-dashboard.ages.at/), 7-day I (15 to 24) = 7 days incidence for SARS-CoV-2 infections per 100,000 inhabitants in Carinthia (AUT) for adolescent aged 15 to 24 years (https://covid19-dashboard.ages.at/), AUT = Austria, P1 = period 1 (September 13, 2021 to October 31, 2021) P2 = period 2 (November 1, 2021 to December 31, 2021), P3 = period 3 (January 1, 2022 to February 28, 2022), P4 = period 4 (March 1, 2022 to April 30, 2022), A = Risk level 1: All students can be tested voluntarily at school for SARS-CoV-2 infection by rapid anterionasal antigen test, B = Risk level 2: Unvaccinated students are tested three times weekly for SARS-CoV-2 infection (twice anterionasal rapid antigen test, once PCR test with mouth rinse) and Students are required to wear a face mask outside of classrooms, PCR = polymerase chain reaction, C = All students are tested three times weekly for SARS-CoV-2 infection (once anterionasal rapid antigen test, twice PCR test with mouth rinse) and Students are required to wear a FFP2 face mask anywhere in school building, FFP2 = filtering face piece generation 2 (FFP2 masks: have a protective effect of > 95 % for airborne particles in controlled laboratory tests; air permeability and airway resistance are standardized. The leakage is < 11 %), D = All students are tested three times weekly for SARS-CoV-2 infection (once anterionasal rapid antigen test, twice PCR test with mouth rinse), B.1.617.2 = Delta variant of SARS-CoV-2 Virus, BA.1 = Omicron variant of SARS-CoV-2 Virus, BA.2/BA.3 = Omicron variant of SARS-CoV-2 Virus.

**eTable 2.** Sample characteristics for total study population and subgroups middle and high school

| Variable | All (n=614) | GB (n=419) | SF (n=195) | **$X^2$** | **t** | ***P* value** | **p-lvl** |
|---|---|---|---|---|---|---|---|
| Female, No. (%) | 265 (43.2%) | 216 (51.6%) | 49 (25.1%) | 37.872 | | <.001 | *** |
| Age at index, mean (SD) | 15.1 ± 2.3 | 15.3 ± 2.3 | 14.8 ± 2.2 | | 2.752 | 0.006 | ** |
| SARS-CoV-2 infection - bevor September 13, 2021, No. (%) | 76 (12.4%) | 44 (10.5%) | 32 (16.4%) | 4.284 | | 0.038 | * |
| SARS-CoV-2 infection - September 13, 2021 to April 30, 2022, No. (%) | 353 (57.5%) | 229 (54.7%) | 124 (63.6%) | 4.348 | | 0.037 | * |
| | M.S. | | | | | | |
| | All (n=289) | GB (n=190) | SF (n=99) | | | | |
| Female, No. (%) | 111 (38.4%) | 84 (44.2%) | 27 (27.3%) | 7.893 | | 0.005 | ** |
| Age at index, mean (SD) | 13.1 ± 1.3 | 13.2 ± 1.3 | 12.9 ± 1.2 | | 2.105 | 0.036 | * |
| SARS-CoV-2 infection - bevor September 13, 2021, No. (%) | 27 (9.3%) | 17 (8.9%) | 10 (10.1%) | 0.102 | | 0.75 | |
| SARS-CoV-2 infection - September 13, 2021 to April 30, 2022, No. (%) | 173 (59.9%) | 107 (56.3%) | 66 (66.7%) | 2.902 | | 0.09 | |
| | H.S. | | | | | | |
| | All (n=325) | GB (n=229) | SF (n=96) | | | | |
| Female, No. (%) | 154 (47.4%) | 132 (57.6%) | 22 (22.9%) | 32.716 | | <.001 | *** |
| Age at index, mean (SD) | 16.9 ± 1.2 | 17.0 ± 1.3 | 16.7 ± 1.2 | | 2.258 | 0.025 | * |
| SARS-CoV-2 infection - bevor September 13, 2021, No. (%) | 49 (15.1%) | 27 (11.8%) | 22 (22.9%) | 6.540 | | 0.011 | * |
| SARS-CoV-2 infection - September 13, 2021 to April 30, 2022, No. (%) | 180 (55.4%) | 122 (53.3%) | 58 (60.4%) | 1.396 | | 0.24 | |

SARS-CoV-2 = severe acute respiratory syndrome coronavirus 2, Data are absolute numbers (No.) and percentages (%), $X^2$ = Chi-Square Test value, t = test statistic t-test, p-lvl (P Value level) * = P < 0.05, ** = P < 0.01, *** = P < 0.001, p-lvl = significance level, No. = Number, % = Percentage, M.S. = middle school (Students aged 13.1 ± 1.3 years old), H.S. = 4-year high school (Students aged 16.9 ± 1.2 years old), GB = students in school classes with a general school branch, SF = students in school classes with sport focus, n = Study population.

**eTable 3:** Cumulative percentage of students with a SARS-CoV-2 infection: students in classes with GB vs students in SF classes for subgroups sex.

| Variable | Time period | Boys | | | | | | | Girls | | | | | | |
|---|---|---|---|---|---|---|---|---|---|---|---|---|---|---|---|
| | | All (n=349) | | $X^2$ | p Value [b] | p-lvl | φ | OR (95% CI) | All (n=265) | | $X^2$ | p Value [b] | p-lvl | φ | OR (95% CI) |
| | | GB (n=203) | SF (n=146) | | | | | | GB (n=216) | SF (n=49) | | | | | |
| CU% SARS-CoV-2 …, No. [%] | …Sep.21 | 1 (0.5%) | 2 (1.4%) | 0.767[a] | 0.57 | | 0.05 | 2.81 (0.25 to 31.24) | 0 (0.0%) | 0 (0.0%) | INC | | | | |
| | …Oct.21 | 3 (1.5%) | 5 (3.4%) | 1.437[a] | 0.29 | | 0.06 | 2.36 (0.56 to 10.05) | 2 (0.9%) | 0 (0.0%) | 0.457[a] | >.99 | | -0.04 | 0.81 (0.77 to 0.86) |
| | …Nov.21 | 10 (4.9%) | 23 (15.8%) | 11.628 | 0.001 | ** | 0.18 | 3.61 (1.66 to 7.84) | 16 (7.4%) | 6 (12.2%) | 1.228[a] | 0.26 | | 0.07 | 1.74 (0.65 to 4.72) |
| | …Dec.21 | 15 (7.4%) | 25 (17.1%) | 7.930 | 0.005 | ** | 0.15 | 2.59 (1.31 to 5.11) | 23 (10.6%) | 7 (14.3%) | 0.526 | 0.47 | | 0.05 | 1.40 (0.56 to 3.47) |
| | …Jan.22 | 35 (17.2%) | 56 (38.4%) | 19.642 | <.001 | *** | 0.24 | 2.99 (1.82 to 4.89) | 46 (21.3%) | 29 (59.2%) | 28.253 | <.001 | *** | 0.33 | 5.36 (2.78 to 10.33) |
| | …Feb.22 | 66 (32.5%) | 83 (56.8%) | 20.559 | <.001 | *** | 0.24 | 2.74 (1.76 to 4.25) | 83 (38.4%) | 32 (65.3%) | 11.748 | 0.001 | ** | 0.21 | 3.02 (1.58 to 5.77) |
| | …Mar.22 | 98 (48.3%) | 87 (59.6%) | 4.363 | 0.037 | * | 0.11 | 1.58 (1.03 to 2.43) | 117 (54.2%) | 33 (67.3%) | 2.825 | 0.09 | | 0.10 | 1.75 (0.91 to 3.36) |
| | …Apr.22 | 104 (51.2%) | 91 (62.3%) | 4.242 | 0.039 | * | 0.11 | 1.58 (1.02 to 2.43) | 125 (57.9%) | 33 (67.3%) | 1.49 | 0.22 | | 0.08 | 1.45 (1.02 to 2.06) |
| | | M.S. (n=178) | | | | | | | M.S. (n=111) | | | | | | |
| | | GB (n=106) | SF (n=72) | | | | | | GB (n=84) | SF (n=27) | | | | | |
| CU% SARS-CoV-2 …, No. [%] | …Sep.21 | 0 (0.0%) | 1 (1.4%) | 1.481[a] | 0.40 | | 0.09 | 0.40 (0.34 to 0.48) | 0 (0.0%) | 0 (0.0%) | INC | | | | |
| | …Oct.21 | 1 (0.9%) | 2 (2.8%) | 0.871[a] | 0.57 | | 0.07 | 3.00 (0.27 to 33.72) | 1 (1.2%) | 0 (0.0%) | 0.324[a] | >.99 | | -0.05 | 0.76 (0.68 to 0.84) |
| | …Nov.21 | 5 (4.7%) | 12 (16.7%) | 7.088 | 0.008 | ** | 0.20 | 4.04 (1.36 to 12.03) | 4 (4.8%) | 3 (11.1%) | 1.394 a | 0.36 | | 0.11 | 2.50 (0.52 to 11.96) |
| | …Dec.21 | 8 (7.5%) | 14 (19.4%) | 5.603 | 0.018 | * | 0.18 | 2.96 (1.17 to 7.48) | 7 (8.3%) | 4 (14.8%) | 0.961 a | 0.46 | | 0.09 | 1.91 (0.51 to 7.12) |
| | …Jan.22 | 17 (16.0%) | 23 (31.9%) | 6.227 | 0.013 | * | 0.19 | 2.46 (1.20 to 5.04) | 16 (19.0%) | 17 (63.0%) | 18.862 | <.001 | *** | 0.41 | 7.23 (2.79 to 18.72) |
| | …Feb.22 | 39 (36.8%) | 40 (55.6%) | 6.115 | 0.013 | * | 0.19 | 2.15 (1.17 to 3.95) | 31 (36.9%) | 20 (74.1%) | 11.366 | 0.001 | ** | 0.32 | 4.89 (1.86 to 12.86) |
| | …Mar.22 | 55 (51.9%) | 43 (59.7%) | 1.064 | 0.30 | | 0.08 | 1.38 (0.75 to 2.52) | 46 (54.8%) | 21 (77.8%) | 4.524 | 0.033 | * | 0.20 | 2.89 (1.06 to 7.89) |
| | …Apr.22 | 57 (53.8%) | 45 (62.5%) | 1.334 | 0.25 | | 0.09 | 1.43 (0.78 to 2.64) | 50 (59.5%) | 21 (77.8%) | 2.954 | 0.09 | | 0.16 | 2.38 (0.87 to 6.51) |
| | | H.S. (n=171) | | | | | | | H.S. (n=154) | | | | | | |
| | | GB (n=97) | SF (n=74) | | | | | | GB (n=132) | SF (n=22) | | | | | |
| CU% SARS-CoV-2 …, No. [%] | …Sep.21 | 1 (1.0%) | 1 (1.4%) | 0.037[a] | >.99 | | 0.02 | 1.32 (0.08 to 21.38) | 0 (0.0%) | 0 (0.0%) | INC | | | | |
| | …Oct.21 | 2 (2.1%) | 3 (4.1%) | 0.587[a] | 0.65 | | 0.06 | 2.00 (0.33 to 12.33) | 1 (1.2%) | 0 (0.0%) | 0.168[a] | >.99 | | -0.03 | 0.86 (0.80 to 0.91) |
| | …Nov.21 | 5 (5.2%) | 11 (14.9%) | 4.667 | 0.031 | * | 0.17 | 3.21 (1.06 to 9.70) | 12 (9.1%) | 3 (13.6%) | 0.443[a] | 0.45 | | 0.05 | 1.58 (0.41 to 6.12) |
| | …Dec.21 | 7 (7.2%) | 11 (14.9%) | 2.607 | 0.11 | | 0.12 | 2.25 (0.83 to 6.11) | 16 (12.1%) | 3 (13.6%) | 0.040[a] | 0.74 | | 0.02 | 1.15 (0.30 to 4.31) |
| | …Jan.22 | 18 (18.6%) | 33 (44.6%) | 13.598 | <.001 | *** | 0.28 | 3.53 (1.78 to 7.02) | 30 (22.7%) | 12 (54.5%) | 9.625 | 0.002 | ** | 0.25 | 4.08 (1.61 to 10.37) |
| | …Feb.22 | 27 (27.8%) | 43 (58.1%) | 15.911 | <.001 | *** | 0.31 | 3.60 (1.90 to 6.82) | 52 (39.4%) | 12 (54.5%) | 1.782 | 0.18 | | 0.11 | 1.85 (0.74 to 4.58) |
| | …Mar.22 | 43 (44.3%) | 44 (59.5%) | 3.845 | 0.05 | * | 0.15 | 1.84 (1.00 to 3.40) | 71 (53.8%) | 12 (54.5%) | 0.004 | 0.95 | | 0.01 | 1.03 (0.42 to 2.55) |
| | …Apr.22 | 47 (48.5%) | 46 (62.2%) | 3.180 | 0.08 | | 0.14 | 1.75 (0.94 to 3.24) | 75 (56.8%) | 12 (54.5%) | 0.040 | 0.84 | | -0.02 | 0.91 (0.37 to 2.26) |

a = 2 cells (50,0%) have expected count less than 5; b = If ( a ) occurred, Fisher's Exact Test was used.

Data are No. (%), n = Study population, GB = students in school classes with a general school branch, SF = students in school classes with sport focus, $X^2$ = Chi-Square Test value, p-lvl (P Value level) * = P < 0.05, ** = P < 0.01, *** = P < 0.001, p-lvl = significance level, φ = Effect size Phi, OR = odds ratio (with 1 for students in classes without sport focus), CI = confidence interval, CU% SARS-CoV-2 = cumulative percentage of students with a SARS-COV-2 infections at the End of…, SARS-CoV-2 = severe acute respiratory syndrome coronavirus 2, P1 = period 1 (September 13, 2021 to October 31, 2021) P2 = period 2 (November 1, 2021 to December 31, 2021), P3 = period 3 (January 1, 2022 to February 28, 2022), P4 = period 4 (March 1, 2022 to April 30, 2022), INC = insufficient number of potential infection cases.

**eTable 4.** Detailed increase of cumulative numbers and percentage of students with a SARS-CoV-2 infection for P1 (Sep.21 and Oct.21).

| Mo. | M.S. | | | | H.S. | | | | Mo. | M.S. | | | | H.S. | | | |
|---|---|---|---|---|---|---|---|---|---|---|---|---|---|---|---|---|---|
| Sep.21 | GB (n=190) | | SF (n=99) | | GB (n=229) | | SF (n=96) | | Oct.21 | GB (n=190) | | SF (n=99) | | GB (n=229) | | SF (n=96) | |
| | No. | % | No. | % | No. | % | No. | % | | No. | % | No. | % | No. | % | No. | % |
| 1 | | | | | | | | | 1 | 0 | 0.00% | 1 | 1.00% | 1 | 0.40% | 1 | 1.00% |
| 2 | | | | | | | | | 2 | 0 | 0.00% | 1 | 1.00% | 1 | 0.40% | 1 | 1.00% |
| 3 | | | | | | | | | 3 | 0 | 0.00% | 1 | 1.00% | 1 | 0.40% | 1 | 1.00% |
| 4 | | | | | | | | | 4 | 0 | 0.00% | 1 | 1.00% | 1 | 0.40% | 1 | 1.00% |
| 5 | | | | | | | | | 5 | 0 | 0.00% | 1 | 1.00% | 2 | 0.90% | 1 | 1.00% |
| 6 | | | | | | | | | 6 | 0 | 0.00% | 1 | 1.00% | 2 | 0.90% | 1 | 1.00% |
| 7 | | | | | | | | | 7 | 0 | 0.00% | 1 | 1.00% | 2 | 0.90% | 1 | 1.00% |
| 8 | | | | | | | | | 8 | 0 | 0.00% | 1 | 1.00% | 2 | 0.90% | 1 | 1.00% |
| 9 | | | | | | | | | 9 | 0 | 0.00% | 1 | 1.00% | 2 | 0.90% | 1 | 1.00% |
| 10 | | | | | | | | | 10 | 0 | 0.00% | 1 | 1.00% | 2 | 0.90% | 1 | 1.00% |
| 11 | | | | | | | | | 11 | 0 | 0.00% | 1 | 1.00% | 2 | 0.90% | 1 | 1.00% |
| 12 | | | | | | | | | 12 | 0 | 0.00% | 1 | 1.00% | 2 | 0.90% | 1 | 1.00% |
| 13 | 0 | 0.00% | 0 | 0.00% | 0 | 0.00% | 0 | 0.00% | 13 | 0 | 0.00% | 1 | 1.00% | 2 | 0.90% | 1 | 1.00% |
| 14 | 0 | 0.00% | 0 | 0.00% | 0 | 0.00% | 0 | 0.00% | 14 | 0 | 0.00% | 1 | 1.00% | 2 | 0.90% | 2 | 2.10% |
| 15 | 0 | 0.00% | 0 | 0.00% | 0 | 0.00% | 0 | 0.00% | 15 | 0 | 0.00% | 1 | 1.00% | 2 | 0.90% | 2 | 2.10% |
| 16 | 0 | 0.00% | 0 | 0.00% | 0 | 0.00% | 0 | 0.00% | 16 | 0 | 0.00% | 1 | 1.00% | 2 | 0.90% | 2 | 2.10% |
| 17 | 0 | 0.00% | 0 | 0.00% | 0 | 0.00% | 0 | 0.00% | 17 | 0 | 0.00% | 1 | 1.00% | 2 | 0.90% | 2 | 2.10% |
| 18 | 0 | 0.00% | 0 | 0.00% | 0 | 0.00% | 0 | 0.00% | 18 | 0 | 0.00% | 1 | 1.00% | 2 | 0.90% | 2 | 2.10% |
| 19 | 0 | 0.00% | 0 | 0.00% | 0 | 0.00% | 0 | 0.00% | 19 | 0 | 0.00% | 1 | 1.00% | 2 | 0.90% | 2 | 2.10% |
| 20 | 0 | 0.00% | 0 | 0.00% | 0 | 0.00% | 0 | 0.00% | 20 | 1 | 0.50% | 2 | 2.00% | 2 | 0.90% | 2 | 2.10% |
| 21 | 0 | 0.00% | 0 | 0.00% | 0 | 0.00% | 0 | 0.00% | 21 | 1 | 0.50% | 2 | 2.00% | 2 | 0.90% | 2 | 2.10% |
| 22 | 0 | 0.00% | 0 | 0.00% | 0 | 0.00% | 0 | 0.00% | 22 | 2 | 1.10% | 2 | 2.00% | 2 | 0.90% | 2 | 2.10% |
| 23 | 0 | 0.00% | 0 | 0.00% | 0 | 0.00% | 1 | 1.00% | 23 | 2 | 1.10% | 2 | 2.00% | 2 | 0.90% | 2 | 2.10% |
| 24 | 0 | 0.00% | 0 | 0.00% | 0 | 0.00% | 1 | 1.00% | 24 | 2 | 1.10% | 2 | 2.00% | 2 | 0.90% | 2 | 2.10% |
| 25 | 0 | 0.00% | 0 | 0.00% | 0 | 0.00% | 1 | 1.00% | 25 | 2 | 1.10% | 2 | 2.00% | 2 | 0.90% | 2 | 2.10% |
| 26 | 0 | 0.00% | 0 | 0.00% | 0 | 0.00% | 1 | 1.00% | 26 | 2 | 1.10% | 2 | 2.00% | 2 | 0.90% | 2 | 2.10% |
| 27 | 0 | 0.00% | 0 | 0.00% | 1 | 0.40% | 1 | 1.00% | 27 | 2 | 1.10% | 2 | 2.00% | 2 | 0.90% | 3 | 3.10% |
| 28 | 0 | 0.00% | 0 | 0.00% | 1 | 0.40% | 1 | 1.00% | 28 | 2 | 1.10% | 2 | 2.00% | 3 | 1.30% | 3 | 3.10% |
| 29 | 0 | 0.00% | 1 | 1.00% | 1 | 0.40% | 1 | 1.00% | 29 | 2 | 1.10% | 2 | 2.00% | 3 | 1.30% | 3 | 3.10% |
| 30 | 0 | 0.00% | 1 | 1.00% | 1 | 0.40% | 1 | 1.00% | 30 | 2 | 1.10% | 2 | 2.00% | 3 | 1.30% | 3 | 3.10% |
| | | | | | | | | | 31 | 2 | 1.10% | 2 | 2.00% | 3 | 1.30% | 3 | 3.10% |

SARS-CoV-2 = severe acute respiratory syndrome coronavirus 2, P1 = period 1 (September 13, 2021 to October 31, 2021), Data are cumulative numbers (No.) and percentages (%) of SARS-CoV-2 infections detected, No. = Number, % = Percentage, Mo. = month, M.S. = middle school (Students aged 13.1 ± 1.3 years old), H.S. = 4-year high school (Students aged 16.9 ± 1.2 years old), GB = students in school classes with a general school branch, SF = students in school classes with sport focus, n = Study population, Sep.21 = September 2021, Oct.21 = October 2021.

**eTable 5.** Detailed increase of cumulative numbers and percentage of students with a SARS-CoV-2 infection for P2 (Nov.21 and Dec.21).

| Mo. | M.S. | | | | H.S. | | | | Mo. | M.S. | | | | H.S. | | | |
|---|---|---|---|---|---|---|---|---|---|---|---|---|---|---|---|---|---|
| Nov.21 | GB (n=190) | | SF (n=99) | | GB (n=229) | | SF (n=96) | | Dec.21 | GB (n=190) | | SF (n=99) | | GB (n=229) | | SF (n=96) | |
| | No. | % | No. | % | No. | % | No. | % | | No. | % | No. | % | No. | % | No. | % |
| 1 | 2 | 1.1% | 3 | 3.0% | 3 | 1.3% | 3 | 3.1% | 1 | 10 | 5.3% | 15 | 15.2% | 17 | 7.4% | 14 | 14.6% |
| 2 | 2 | 1.1% | 4 | 4.0% | 3 | 1.3% | 3 | 3.1% | 2 | 10 | 5.3% | 15 | 15.2% | 17 | 7.4% | 14 | 14.6% |
| 3 | 2 | 1.1% | 5 | 5.1% | 3 | 1.3% | 4 | 4.2% | 3 | 10 | 5.3% | 15 | 15.2% | 17 | 7.4% | 14 | 14.6% |
| 4 | 3 | 1.6% | 5 | 5.1% | 4 | 1.7% | 4 | 4.2% | 4 | 10 | 5.3% | 15 | 15.2% | 17 | 7.4% | 14 | 14.6% |
| 5 | 3 | 1.6% | 5 | 5.1% | 6 | 2.6% | 4 | 4.2% | 5 | 10 | 5.3% | 15 | 15.2% | 18 | 7.9% | 14 | 14.6% |
| 6 | 3 | 1.6% | 5 | 5.1% | 6 | 2.6% | 5 | 5.2% | 6 | 11 | 5.8% | 16 | 16.2 | 19 | 8.3% | 14 | 14.6% |
| 7 | 3 | 1.6% | 5 | 5.1% | 7 | 3.1% | 5 | 5.2% | 7 | 11 | 5.8% | 16 | 16.2% | 20 | 8.7% | 14 | 14.6% |
| 8 | 3 | 1.6% | 6 | 6.1% | 8 | 3.5% | 7 | 7.3% | 8 | 11 | 5.8% | 16 | 16.2% | 20 | 8.7% | 14 | 14.6% |
| 9 | 3 | 1.6% | 6 | 6.1% | 8 | 3.5% | 8 | 8.3% | 9 | 11 | 5.8% | 16 | 16.2% | 20 | 8.7% | 14 | 14.6% |
| 10 | 3 | 1.6% | 6 | 6.1% | 9 | 3.9% | 9 | 9.4% | 10 | 12 | 6.3% | 16 | 16.2% | 21 | 9.2% | 14 | 14.6% |
| 11 | 3 | 1.6% | 6 | 6.1% | 11 | 4.8% | 9 | 9.4% | 11 | 13 | 6.8% | 16 | 16.2% | 21 | 9.2% | 14 | 14.6% |
| 12 | 4 | 2.1% | 6 | 6.1% | 11 | 4.8% | 9 | 9.4% | 12 | 13 | 6.8% | 16 | 16.2% | 21 | 9.2% | 14 | 14.6% |
| 13 | 5 | 2.6% | 6 | 6.1% | 11 | 4.8% | 9 | 9.4% | 13 | 14 | 7.4% | 16 | 16.2% | 23 | 10.0% | 14 | 14.6% |
| 14 | 5 | 2.6% | 6 | 6.1% | 12 | 5.2% | 9 | 9.4% | 14 | 14 | 7.4% | 16 | 16.2% | 23 | 10.0% | 14 | 14.6% |
| 15 | 6 | 3.2% | 6 | 6.1% | 13 | 5.7% | 10 | 10.4% | 15 | 15 | 7.9% | 16 | 16.2% | 23 | 10.0% | 14 | 14.6% |
| 16 | 6 | 3.2% | 7 | 7.1% | 13 | 5.7% | 10 | 10.4% | 16 | 15 | 7.9% | 16 | 16.2% | 23 | 10.0% | 14 | 14.6% |
| 17 | 6 | 3.2% | 7 | 7.1% | 14 | 6.1% | 10 | 10.4% | 17 | 15 | 7.9% | 16 | 16.2% | 23 | 10.0% | 14 | 14.6% |
| 18 | 6 | 3.2% | 7 | 7.1% | 15 | 6.6% | 10 | 10.4% | 18 | 15 | 7.9% | 16 | 16.2% | 23 | 10.0% | 14 | 14.6% |
| 19 | 6 | 3.2% | 7 | 7.1% | 16 | 7.0% | 11 | 11.5% | 19 | 15 | 7.9% | 16 | 16.2% | 23 | 10.0% | 14 | 14.6% |
| 20 | 6 | 3.2% | 7 | 7.1% | 16 | 7.0% | 13 | 13.5% | 20 | 15 | 7.9% | 16 | 16.2% | 23 | 10.0% | 14 | 14.6% |
| 21 | 6 | 3.2% | 7 | 7.1% | 16 | 7.0% | 13 | 13.5% | 21 | 15 | 7.9% | 16 | 16.2 | 23 | 10.0% | 14 | 14.6% |
| 22 | 6 | 3.2% | 7 | 7.1% | 16 | 7.0% | 13 | 13.5% | 22 | 15 | 7.9% | 16 | 16.2% | 23 | 10.0% | 14 | 14.6% |
| 23 | 6 | 3.2% | 7 | 7.1% | 16 | 7.0% | 13 | 13.5% | 23 | 15 | 7.9% | 16 | 16.2% | 23 | 10.0% | 14 | 14.6% |
| 24 | 6 | 3.2% | 9 | 9.1% | 16 | 7.0% | 13 | 13.5% | 24 | 15 | 7.9% | 16 | 16.2% | 23 | 10.0% | 14 | 14.6% |
| 25 | 7 | 3.7% | 12 | 12.1% | 16 | 7.0% | 13 | 13.5% | 25 | 15 | 7.9% | 16 | 16.2% | 23 | 10.0% | 14 | 14.6% |
| 26 | 8 | 4.2% | 14 | 14.1% | 17 | 7.4% | 14 | 14.6% | 26 | 15 | 7.9% | 17 | 17.2% | 23 | 10.0% | 14 | 14.6% |
| 27 | 8 | 4.2% | 14 | 14.1% | 17 | 7.4% | 14 | 14.6% | 27 | 15 | 7.9% | 17 | 17.2% | 23 | 10.0% | 14 | 14.6% |
| 28 | 8 | 4.2% | 14 | 14.1% | 17 | 7.4% | 14 | 14.6% | 28 | 15 | 7.9% | 17 | 17.2% | 23 | 10.0% | 14 | 14.6% |
| 29 | 8 | 4.2% | 15 | 15.2% | 17 | 7.4% | 14 | 14.6% | 29 | 15 | 7.9% | 18 | 18.2% | 23 | 10.0% | 14 | 14.6% |
| 30 | 9 | 4.7% | 15 | 15.2% | 17 | 7.4% | 14 | 14.6% | 30 | 15 | 7.9% | 18 | 18.2% | 23 | 10.0% | 14 | 14.6% |
| | | | | | | | | | 31 | 15 | 7.9% | 18 | 18.2% | 23 | 10.0% | 14 | 14.6% |

SARS-CoV-2 = severe acute respiratory syndrome coronavirus 2, P2 = period 2 (November 1, 2021 to December 31, 2021), Data are cumulative numbers (No.) and percentages (%) of SARS-CoV-2 infections detected, No. = Number, % = Percentage, Mo. = month, M.S. = middle school (Students aged 13.1 ± 1.3 years old), H.S. = 4-year high school (Students aged 16.9 ± 1.2 years old), GB = students in school classes with a general school branch, SF = students in school classes with sport focus, n = Study population, Nov.21 = November 2021, Dec.21 = December 2021.

**eTable 6.** Detailed increase of cumulative numbers and percentage of students with a SARS-CoV-2 infection for P3 (Jan.22 and Feb.22).

| Mo. | M.S. | | | | H.S. | | | | Mo. | M.S. | | | | H.S. | | | |
|---|---|---|---|---|---|---|---|---|---|---|---|---|---|---|---|---|---|
| Jan.22 | GB (n=190) | | SF (n=99) | | GB (n=229) | | SF (n=96) | | Feb.22 | GB (n=190) | | SF (n=99) | | GB (n=229) | | SF (n=96) | |
| | No. | % | No. | % | No. | % | No. | % | | No. | % | No. | % | No. | % | No. | % |
| 1 | 15 | 7.9% | 18 | 18.2% | 24 | 10.5% | 14 | 14.6% | 1 | 36 | 18.9% | 40 | 40.4% | 50 | 21.8% | 46 | 47.9% |
| 2 | 15 | 7.9% | 18 | 18.2% | 24 | 10.5% | 14 | 14.6% | 2 | 37 | 19.5% | 44 | 44.4% | 51 | 22.3% | 46 | 47.9% |
| 3 | 15 | 7.9% | 18 | 18.2% | 24 | 10.5% | 14 | 14.6% | 3 | 39 | 20.5% | 44 | 44.4% | 51 | 22.3% | 48 | 50.0% |
| 4 | 15 | 7.9% | 18 | 18.2% | 24 | 10.5% | 15 | 15.6% | 4 | 41 | 21.6% | 45 | 45.5% | 51 | 22.3% | 49 | 51.0% |
| 5 | 16 | 8.4% | 18 | 18.2% | 24 | 10.5% | 15 | 15.6% | 5 | 42 | 22.1% | 46 | 46.5% | 52 | 22.7% | 50 | 52.1% |
| 6 | 16 | 8.4% | 18 | 18.2% | 24 | 10.5% | 16 | 16.7% | 6 | 42 | 22.1% | 47 | 47.5% | 52 | 22.7% | 52 | 54.2% |
| 7 | 16 | 8.4% | 19 | 19.2% | 25 | 10.9% | 16 | 16.7% | 7 | 42 | 22.1% | 48 | 48.5% | 54 | 23.6% | 52 | 54.2% |
| 8 | 16 | 8.4% | 19 | 19.2% | 26 | 11.4% | 17 | 17.7% | 8 | 43 | 22.6% | 51 | 51.5% | 55 | 24.0% | 53 | 55.2% |
| 9 | 17 | 8.9% | 20 | 20.2% | 27 | 11.8% | 17 | 17.7% | 9 | 45 | 23.7% | 52 | 52.5% | 57 | 24.9% | 53 | 55.2% |
| 10 | 21 | 11.1% | 21 | 21.2% | 27 | 11.8% | 18 | 18.8% | 10 | 47 | 24.7% | 55 | 55.6% | 58 | 25.3% | 54 | 56.3% |
| 11 | 23 | 12.1% | 22 | 22.2% | 27 | 11.8% | 19 | 19.8% | 11 | 47 | 24.7% | 55 | 55.6% | 58 | 25.3% | 54 | 56.3% |
| 12 | 23 | 12.1% | 22 | 22.2% | 27 | 11.8% | 20 | 20.8% | 12 | 49 | 25.8% | 55 | 55.6% | 58 | 25.3% | 54 | 56.3% |
| 13 | 24 | 12.6% | 22 | 22.2% | 27 | 11.8% | 20 | 20.8% | 13 | 51 | 26.8% | 55 | 55.6% | 59 | 25.8% | 54 | 56.3% |
| 14 | 24 | 12.6% | 22 | 22.2% | 27 | 11.8% | 20 | 20.8% | 14 | 52 | 27.4% | 55 | 55.6% | 61 | 26.6% | 54 | 56.3% |
| 15 | 24 | 12.6% | 22 | 22.2% | 30 | 13.1% | 20 | 20.8% | 15 | 52 | 27.4% | 56 | 56.6% | 66 | 28.8% | 54 | 56.3% |
| 16 | 24 | 12.6% | 22 | 22.2% | 30 | 13.1% | 22 | 22.9% | 16 | 52 | 27.4% | 56 | 56.6% | 66 | 28.8% | 54 | 56.3% |
| 17 | 24 | 12.6% | 23 | 23.2% | 33 | 14.4% | 23 | 24.0% | 17 | 55 | 28.9% | 56 | 56.6% | 66 | 28.8% | 54 | 56.3% |
| 18 | 25 | 13.2% | 24 | 24.2% | 34 | 14.8% | 27 | 28.1% | 18 | 56 | 29.5% | 57 | 57.6% | 68 | 29.7% | 54 | 56.3% |
| 19 | 26 | 13.7% | 24 | 24.2% | 37 | 16.2% | 29 | 30.2% | 19 | 59 | 31.1% | 57 | 57.6% | 68 | 29.7% | 54 | 56.3% |
| 20 | 26 | 13.7% | 25 | 25.3% | 39 | 17.0% | 31 | 32.3% | 20 | 63 | 33.2% | 57 | 57.6% | 69 | 30.1% | 54 | 56.3% |
| 21 | 27 | 14.2% | 27 | 27.3% | 40 | 17.5% | 34 | 35.4% | 21 | 65 | 34.2% | 58 | 58.6% | 71 | 31.0% | 54 | 56.3% |
| 22 | 28 | 14.7% | 28 | 28.3% | 40 | 17.5% | 39 | 40.6% | 22 | 67 | 35.3% | 58 | 58.6% | 74 | 32.3% | 55 | 57.3% |
| 23 | 29 | 15.3% | 29 | 29.3% | 40 | 17.5% | 40 | 41.7% | 23 | 67 | 35.3% | 59 | 59.6% | 77 | 33.6% | 55 | 57.3% |
| 24 | 29 | 15.3% | 30 | 30.3% | 40 | 17.5% | 41 | 42.7% | 24 | 67 | 35.3% | 59 | 59.6% | 78 | 34.1% | 55 | 57.3% |
| 25 | 30 | 15.8% | 30 | 30.3% | 41 | 17.9% | 42 | 43.8% | 25 | 69 | 36.3% | 59 | 59.6% | 78 | 34.1% | 55 | 57.3% |
| 26 | 30 | 15.8% | 34 | 34.3% | 41 | 17.9% | 42 | 43.8% | 26 | 69 | 36.3% | 60 | 60.6% | 78 | 34.1% | 55 | 57.3% |
| 27 | 31 | 16.3% | 37 | 37.4% | 42 | 18.3% | 44 | 45.8% | 27 | 69 | 36.3% | 60 | 60.6% | 78 | 34.1% | 55 | 57.3% |
| 28 | 32 | 16.8% | 38 | 38.4% | 42 | 18.3% | 44 | 45.8% | 28 | 70 | 36.8% | 60 | 60.6% | 79 | 34.5% | 55 | 57.3% |
| 29 | 32 | 16.8% | 38 | 38.4% | 43 | 18.8% | 44 | 45.8% | | | | | | | | | |
| 30 | 32 | 16.8% | 38 | 38.4% | 46 | 20.1% | 44 | 45.8% | | | | | | | | | |
| 31 | 33 | 17.4% | 40 | 40.4% | 48 | 21.0% | 45 | 46.9% | | | | | | | | | |

SARS-CoV-2 = severe acute respiratory syndrome coronavirus 2, P3 = period 3 (January 1, 2022 to February 28, 2022), Data are cumulative numbers (No.) and percentages (%) of SARS-CoV-2 infections detected, No. = Number, % = Percentage, Mo. = month, M.S. = middle school (Students aged 13.1 ± 1.3 years old), H.S. = 4-year high school (Students aged 16.9 ± 1.2 years old), GB = students in school classes with a general school branch, SF = students in school classes with sport focus, n = Study population, Jan.22 = January 2022, Feb.22 = February 2022.

**eTable 7.** Detailed increase of cumulative numbers and percentage of students with a SARS-CoV-2 infection for P4 (Mar.22 and Apr.22).

| Mo. | M.S. | | | | H.S. | | | | Mo. | M.S. | | | | H.S. | | | |
|---|---|---|---|---|---|---|---|---|---|---|---|---|---|---|---|---|---|
| Mar.22 | GB (n=190) | | SF (n=99) | | GB (n=229) | | SF (n=96) | | Apr.22 | GB (n=190) | | SF (n=99) | | GB (n=229) | | SF (n=96) | |
| | No. | % | No. | % | No. | % | No. | % | | No. | % | No. | % | No. | % | No. | % |
| 1 | 70 | 36.8% | 61 | 61.6% | 82 | 35.8% | 55 | 57.3% | 1 | 101 | 53.2% | 64 | 64.6% | 115 | 50.2% | 56 | 58.3% |
| 2 | 72 | 37.9% | 61 | 61.6% | 83 | 36.2% | 55 | 57.3% | 2 | 102 | 53.7% | 64 | 64.6% | 115 | 50.2% | 56 | 58.3% |
| 3 | 73 | 38.4% | 61 | 61.6% | 83 | 36.2% | 55 | 57.3% | 3 | 105 | 55.3% | 64 | 64.6% | 115 | 50.2% | 56 | 58.3% |
| 4 | 73 | 38.4% | 62 | 62.6% | 84 | 36.7% | 55 | 57.3% | 4 | 105 | 55.3% | 65 | 65.7% | 115 | 50.2% | 56 | 58.3% |
| 5 | 73 | 38.4% | 62 | 62.6% | 86 | 37.6% | 55 | 57.3% | 5 | 105 | 55.3% | 65 | 65.7% | 115 | 50.2% | 56 | 58.3% |
| 6 | 73 | 38.4% | 62 | 62.6% | 86 | 37.6% | 55 | 57.3% | 6 | 105 | 55.3% | 65 | 65.7% | 116 | 50.7% | 56 | 58.3% |
| 7 | 73 | 38.4% | 62 | 62.6% | 87 | 38.0% | 56 | 58.3% | 7 | 105 | 55.3% | 65 | 65.7% | 117 | 51.1% | 56 | 58.3% |
| 8 | 74 | 38.9% | 62 | 62.6% | 87 | 38.0% | 56 | 58.3% | 8 | 105 | 55.3% | 65 | 65.7% | 118 | 51.5% | 56 | 58.3% |
| 9 | 78 | 41.1% | 62 | 62.6% | 90 | 39.3% | 56 | 58.3% | 9 | 105 | 55.3% | 65 | 65.7% | 118 | 51.5% | 56 | 58.3% |
| 10 | 80 | 42.1% | 62 | 62.6% | 93 | 40.6% | 56 | 58.3% | 10 | 106 | 55.8% | 65 | 65.7% | 118 | 51.5% | 56 | 58.3% |
| 11 | 83 | 43.7% | 62 | 62.6% | 93 | 40.6% | 56 | 58.3% | 11 | 106 | 55.8% | 65 | 65.7% | 118 | 51.5% | 56 | 58.3% |
| 12 | 86 | 45.3% | 62 | 62.6% | 93 | 40.6% | 56 | 58.3% | 12 | 106 | 55.8% | 65 | 65.7% | 118 | 51.5% | 56 | 58.3% |
| 13 | 87 | 45.8% | 62 | 62.6% | 93 | 40.6% | 56 | 58.3% | 13 | 106 | 55.8% | 65 | 65.7% | 118 | 51.5% | 56 | 58.3% |
| 14 | 89 | 46.8% | 62 | 62.6% | 93 | 40.6% | 56 | 58.3% | 14 | 106 | 55.8% | 65 | 65.7% | 119 | 52.0% | 56 | 58.3% |
| 15 | 91 | 47.9% | 62 | 62.6% | 94 | 41.0% | 56 | 58.3% | 15 | 107 | 56.3% | 65 | 65.7% | 119 | 52.0% | 56 | 58.3% |
| 16 | 95 | 50.0% | 62 | 62.6% | 98 | 42.8% | 56 | 58.3% | 16 | 107 | 56.3% | 65 | 65.7% | 119 | 52.0% | 56 | 58.3% |
| 17 | 96 | 50.5% | 62 | 62.6% | 99 | 43.2% | 56 | 58.3% | 17 | 107 | 56.3% | 65 | 65.7% | 119 | 52.0% | 57 | 59.4% |
| 18 | 96 | 50.5% | 62 | 62.6% | 101 | 44.1% | 56 | 58.3% | 18 | 107 | 56.3% | 65 | 65.7% | 119 | 52.0% | 57 | 59.4% |
| 19 | 96 | 50.5% | 62 | 62.6% | 101 | 44.1% | 56 | 58.3% | 19 | 107 | 56.3% | 65 | 65.7% | 120 | 52.4% | 57 | 59.4% |
| 20 | 96 | 50.5% | 62 | 62.6% | 105 | 45.9% | 56 | 58.3% | 20 | 107 | 56.3% | 65 | 65.7% | 121 | 52.8% | 57 | 59.4% |
| 21 | 97 | 51.1% | 62 | 62.6% | 107 | 46.7% | 56 | 58.3% | 21 | 107 | 56.3% | 65 | 65.7% | 121 | 52.8% | 57 | 59.4% |
| 22 | 98 | 51.6% | 62 | 62.6% | 110 | 48.0% | 56 | 58.3% | 22 | 107 | 56.3% | 65 | 65.7% | 121 | 52.8% | 57 | 59.4% |
| 23 | 98 | 51.6% | 63 | 63.6% | 112 | 48.9% | 56 | 58.3% | 23 | 107 | 56.3% | 65 | 65.7% | 121 | 52.8% | 57 | 59.4% |
| 24 | 99 | 52.1% | 63 | 63.6% | 112 | 48.9% | 56 | 58.3% | 24 | 107 | 56.3% | 65 | 65.7% | 121 | 52.8% | 57 | 59.4% |
| 25 | 99 | 52.1% | 63 | 63.6% | 112 | 48.9% | 56 | 58.3% | 25 | 107 | 56.3% | 65 | 65.7% | 122 | 53.3% | 57 | 59.4% |
| 26 | 100 | 52.6% | 63 | 63.6% | 112 | 48.9% | 56 | 58.3% | 26 | 107 | 56.3% | 65 | 65.7% | 122 | 53.3% | 58 | 60.4% |
| 27 | 100 | 52.6% | 63 | 63.6% | 112 | 48.9% | 56 | 58.3% | 27 | 107 | 56.3% | 65 | 65.7% | 122 | 53.3% | 58 | 60.4% |
| 28 | 101 | 53.2% | 63 | 63.6% | 114 | 49.8% | 56 | 58.3% | 28 | 107 | 56.3% | 65 | 65.7% | 122 | 53.3% | 58 | 60.4% |
| 29 | 101 | 53.2% | 64 | 64.6% | 114 | 49.8% | 56 | 58.3% | 29 | 107 | 56.3% | 65 | 65.7% | 122 | 53.3% | 58 | 60.4% |
| 30 | 101 | 53.2% | 64 | 64.6% | 114 | 49.8% | 56 | 58.3% | 30 | 107 | 56.3% | 66 | 66.7% | 122 | 53.3% | 58 | 60.4% |
| 31 | 101 | 53.2% | 64 | 64.6% | 114 | 49.8% | 56 | 58.3% | | | | | | | | | |

SARS-CoV-2 = severe acute respiratory syndrome coronavirus 2, , P4 = period 4 (March 1, 2022 to April 30, 2022), Data are cumulative numbers (No.) and percentages (%) of SARS-CoV-2 infections detected, No. = Number, % = Percentage, Mo. = month, M.S. = middle school (Students aged 13.1 ± 1.3 years old), H.S. = 4-year high school (Students aged 16.9 ± 1.2 years old), GB = students in school classes with a general school branch, SF = students in school classes with sport focus, n = Study population, Mar.22 = March 2022, Apr.22 = April 2022.

**eTable 8.** Detailed 7-day incidences for detected SARS-CoV-2 infections for P1 (Sep.21 and Oct.21).

| Mo. | M.S. | | H.S. | | AGES | | Mo. | M.S. | | H.S. | | AGES | |
|---|---|---|---|---|---|---|---|---|---|---|---|---|---|
| Sep.21 | GB (n=190) | SF (n=99) | GB (n=229) | SF (n=96) | I | II | Oct.21 | GB (n=190) | SF (n=99) | GB (n=229) | SF (n=96) | I | II |
| 1 | | | | | | | 1 | 0.0 | 1010.1 | 436.7 | 0.0 | 422.5 | 197.9 |
| 2 | | | | | | | 2 | 0.0 | 1010.1 | 436.7 | 0.0 | 410.7 | 203.4 |
| 3 | | | | | | | 3 | 0.0 | 1010.1 | 436.7 | 0.0 | 383.0 | 186.8 |
| 4 | | | | | | | 4 | 0.0 | 1010.1 | 0.0 | 0.0 | 363.3 | 172.0 |
| 5 | | | | | | | 5 | 0.0 | 1010.1 | 436.7 | 0.0 | 353.4 | 209.0 |
| 6 | | | | | | | 6 | 0.0 | 0.0 | 436.7 | 0.0 | 317.9 | 212.7 |
| 7 | | | | | | | 7 | 0.0 | 0.0 | 436.7 | 0.0 | 302.1 | 214.5 |
| 8 | | | | | | | 8 | 0.0 | 0.0 | 436.7 | 0.0 | 306.0 | 216.4 |
| 9 | | | | | | | 9 | 0.0 | 0.0 | 436.7 | 0.0 | 300.1 | 223.8 |
| 10 | | | | | | | 10 | 0.0 | 0.0 | 436.7 | 0.0 | 296.2 | 225.6 |
| 11 | | | | | | | 11 | 0.0 | 0.0 | 436.7 | 0.0 | 276.4 | 242.3 |
| 12 | | | | | | | 12 | 0.0 | 0.0 | 0.0 | 0.0 | 246.8 | 227.5 |
| 13 | 0.0 | 0.0 | 0.0 | 0.0 | 207.3 | 183.1 | 13 | 0.0 | 0.0 | 0.0 | 0.0 | 270.5 | 231.2 |
| 14 | 0.0 | 0.0 | 0.0 | 0.0 | 215.2 | 194.2 | 14 | 0.0 | 0.0 | 0.0 | 1041.7 | 266.5 | 240.4 |
| 15 | 0.0 | 0.0 | 0.0 | 0.0 | 256.7 | 214.5 | 15 | 0.0 | 0.0 | 0.0 | 1041.7 | 258.6 | 255.2 |
| 16 | 0.0 | 0.0 | 0.0 | 0.0 | 229.0 | 203.4 | 16 | 0.0 | 0.0 | 0.0 | 1041.7 | 246.8 | 260.8 |
| 17 | 0.0 | 0.0 | 0.0 | 0.0 | 240.9 | 210.8 | 17 | 0.0 | 0.0 | 0.0 | 1041.7 | 256.7 | 273.7 |
| 18 | 0.0 | 0.0 | 0.0 | 0.0 | 225.1 | 197.9 | 18 | 0.0 | 0.0 | 0.0 | 1041.7 | 284.3 | 275.6 |
| 19 | 0.0 | 0.0 | 0.0 | 0.0 | 211.3 | 184.9 | 19 | 0.0 | 0.0 | 0.0 | 1041.7 | 329.7 | 308.9 |
| 20 | 0.0 | 0.0 | 0.0 | 0.0 | 191.5 | 168.3 | 20 | 526.3 | 1010.1 | 436.7 | 1041.7 | 371.2 | 353.2 |
| 21 | 0.0 | 0.0 | 0.0 | 0.0 | 211.3 | 166.4 | 21 | 526.3 | 1010.1 | 436.7 | 0.0 | 402.8 | 336.6 |
| 22 | 0.0 | 0.0 | 0.0 | 0.0 | 201.4 | 144.3 | 22 | 1052.6 | 1010.1 | 436.7 | 0.0 | 438.3 | 371.7 |
| 23 | 0.0 | 0.0 | 0.0 | 1041.7 | 223.1 | 138.7 | 23 | 1052.6 | 1010.1 | 436.7 | 0.0 | 535.1 | 416.1 |
| 24 | 0.0 | 0.0 | 0.0 | 1041.7 | 217.2 | 135.0 | 24 | 1052.6 | 1010.1 | 436.7 | 0.0 | 556.8 | 419.8 |
| 25 | 0.0 | 0.0 | 0.0 | 1041.7 | 258.6 | 138.7 | 25 | 1052.6 | 1010.1 | 436.7 | 0.0 | 645.6 | 442.0 |
| 26 | 0.0 | 0.0 | 0.0 | 1041.7 | 294.2 | 162.7 | 26 | 1052.6 | 1010.1 | 436.7 | 0.0 | 695.0 | 445.7 |
| 27 | 0.0 | 0.0 | 436.7 | 1041.7 | 337.6 | 184.9 | 27 | 526.3 | 0.0 | 0.0 | 1041.7 | 671.3 | 455.0 |
| 28 | 0.0 | 0.0 | 436.7 | 1041.7 | 367.2 | 184.9 | 28 | 526.3 | 0.0 | 436.7 | 1041.7 | 750.3 | 543.7 |
| 29 | 0.0 | 1010.1 | 436.7 | 1041.7 | 400.8 | 199.7 | 29 | 0.0 | 0.0 | 436.7 | 1041.7 | 825.3 | 580.7 |
| 30 | 0.0 | 1010.1 | 436.7 | 0.0 | 422.5 | 203.4 | 30 | 0.0 | 0.0 | 436.7 | 1041.7 | 811.5 | 601.1 |
| | | | | | | | 31 | 0.0 | 0.0 | 436.7 | 1041.7 | 825.3 | 615.9 |

SARS-CoV-2 = severe acute respiratory syndrome coronavirus 2, P1 = period 1 (September 13, 2021 to October 31, 2021), Data are raw daily 7-day incidences extrapolated to 100,000 persons, Mo. = month, M.S. = middle school (students aged 13.1 ± 1.3 years old), H.S. = 4-year high school (students aged 16.9 ± 1.2 years old), AGES = Austrian Agency for Health and Food Safety, GB = students in school classes with a general school branch, SF = students in school classes with sport focus, I = 7 days incidence for SARS-CoV-2 infections per 100,000 inhabitants in Carinthia (AUT) for children aged 5 to 14 years (https://covid19-dashboard.ages.at/), II = 7 days incidence for SARS-CoV-2 infections per 100,000 inhabitants in Carinthia (AUT) for adolescent aged 15 to 24 years (https://covid19-dashboard.ages.at/), AUT = Austria, n = Study population, Sep.21 = September 2021, Oct.21 = October 2021.

**eTable 9.** Detailed 7-day incidences for detected SARS-CoV-2 infections for P2 (Nov.21 and Dec.21).

| Mo. | M.S. | | H.S. | | AGES | | Mo. | M.S. | | H.S. | | AGES | |
|---|---|---|---|---|---|---|---|---|---|---|---|---|---|
| Nov.21 | GB (n=190) | SF (n=99) | GB (n=229) | SF (n=96) | I | II | Dec.21 | GB (n=190) | SF (n=99) | GB (n=229) | SF (n=96) | I | II |
| 1 | 0.0 | 1010.1 | 436.7 | 1041.7 | 813.4 | 630.7 | 1 | 2105.3 | 6060.6 | 436.7 | 1041.7 | 3161.0 | 1387.1 |
| 2 | 0.0 | 2020.2 | 436.7 | 1041.7 | 795.7 | 634.4 | 2 | 1579.0 | 3030.3 | 436.7 | 1041.7 | 2657.5 | 1209.5 |
| 3 | 0.0 | 3030.3 | 436.7 | 1041.7 | 801.6 | 686.1 | 3 | 1052.6 | 1010.1 | 0.0 | 0.0 | 2221.2 | 1070.8 |
| 4 | 526.3 | 3030.3 | 436.7 | 1041.7 | 894.4 | 828.5 | 4 | 1052.6 | 1010.1 | 0.0 | 0.0 | 1842.1 | 934.0 |
| 5 | 526.3 | 3030.3 | 1310.0 | 2083.3 | 943.8 | 941.4 | 5 | 1052.6 | 1010.1 | 436.7 | 0.0 | 1642.7 | 832.2 |
| 6 | 526.3 | 3030.3 | 1310.0 | 3125.0 | 937.8 | 1050.5 | 6 | 1579.0 | 1010.1 | 873.4 | 0.0 | 1453.1 | 786.0 |
| 7 | 526.3 | 3030.3 | 1746.7 | 3125.0 | 959.5 | 1142.9 | 7 | 1052.6 | 1010.1 | 1310.0 | 0.0 | 1230.0 | 686.1 |
| 8 | 526.3 | 3030.3 | 2183.4 | 4166.7 | 973.4 | 1274.3 | 8 | 526.3 | 1010.1 | 1310.0 | 0.0 | 937.8 | 619.6 |
| 9 | 526.3 | 2020.2 | 2183.4 | 5208.3 | 1066.2 | 1355.6 | 9 | 526.3 | 1010.1 | 1310.0 | 0.0 | 764.1 | 541.9 |
| 10 | 526.3 | 1010.1 | 2620.1 | 5208.3 | 1551.9 | 1644.1 | 10 | 1052.6 | 1010.1 | 1746.7 | 0.0 | 856.9 | 562.2 |
| 11 | 0.0 | 1010.1 | 3056.8 | 5208.3 | 1555.8 | 1614.5 | 11 | 1579.0 | 1010.1 | 1746.7 | 0.0 | 740.4 | 525.2 |
| 12 | 526.3 | 1010.1 | 2183.4 | 4166.7 | 1561.7 | 1614.5 | 12 | 1579.0 | 1010.1 | 1310.0 | 0.0 | 738.4 | 530.8 |
| 13 | 1052.6 | 1010.1 | 2183.4 | 3125.0 | 1660.4 | 1551.7 | 13 | 1579.0 | 0.0 | 1746.7 | 0.0 | 661.4 | 471.6 |
| 14 | 1052.6 | 1010.1 | 2183.4 | 3125.0 | 1737.4 | 1548.0 | 14 | 1579.0 | 0.0 | 1310.0 | 0.0 | 576.5 | 408.7 |
| 15 | 1579.0 | 0.0 | 2183.4 | 3125.0 | 1891.4 | 1549.8 | 15 | 2105.3 | 0.0 | 1310.0 | 0.0 | 616.0 | 379.1 |
| 16 | 1579.0 | 1010.1 | 2183.4 | 2083.3 | 2084.9 | 1670.0 | 16 | 2105.3 | 0.0 | 1310.0 | 0.0 | 647.6 | 384.7 |
| 17 | 1579.0 | 1010.1 | 2183.4 | 1041.7 | 2460.1 | 1651.5 | 17 | 1579.0 | 0.0 | 873.4 | 0.0 | 501.5 | 329.2 |
| 18 | 1579.0 | 1010.1 | 1746.7 | 1041.7 | 2851.0 | 1832.8 | 18 | 1052.6 | 0.0 | 873.4 | 0.0 | 493.6 | 295.9 |
| 19 | 1052.6 | 1010.1 | 2183.4 | 2083.3 | 3054.4 | 1890.1 | 19 | 1052.6 | 0.0 | 873.4 | 0.0 | 466.0 | 260.8 |
| 20 | 526.3 | 1010.1 | 2183.4 | 4166.7 | 3324.8 | 2051.0 | 20 | 526.3 | 0.0 | 0.0 | 0.0 | 454.1 | 233.0 |
| 21 | 526.3 | 1010.1 | 1746.7 | 4166.7 | 3340.6 | 1997.4 | 21 | 526.3 | 0.0 | 0.0 | 0.0 | 446.2 | 227.5 |
| 22 | 0.0 | 1010.1 | 1310.0 | 3125.0 | 3372.2 | 1986.3 | 22 | 0.0 | 0.0 | 0.0 | 0.0 | 430.4 | 197.9 |
| 23 | 0.0 | 0.0 | 1310.0 | 3125.0 | 3295.2 | 1871.6 | 23 | 0.0 | 0.0 | 0.0 | 0.0 | 438.3 | 188.6 |
| 24 | 0.0 | 3030.3 | 873.4 | 3125.0 | 3228.1 | 1786.5 | 24 | 0.0 | 0.0 | 0.0 | 0.0 | 394.9 | 157.2 |
| 25 | 526.3 | 6060.6 | 436.7 | 3125.0 | 3279.4 | 1653.4 | 25 | 0.0 | 0.0 | 0.0 | 0.0 | 359.3 | 146.1 |
| 26 | 1052.6 | 8080.8 | 436.7 | 3125.0 | 3435.4 | 1585.0 | 26 | 0.0 | 1010.1 | 0.0 | 0.0 | 357.4 | 166.4 |
| 27 | 1052.6 | 8080.8 | 436.7 | 1041.7 | 3514.4 | 1505.4 | 27 | 0.0 | 1010.1 | 0.0 | 0.0 | 359.3 | 172.0 |
| 28 | 1052.6 | 8080.8 | 436.7 | 1041.7 | 3609.2 | 1572.0 | 28 | 0.0 | 1010.1 | 0.0 | 0.0 | 333.7 | 186.8 |
| 29 | 1052.6 | 9090.9 | 436.7 | 1041.7 | 3605.2 | 1527.6 | 29 | 0.0 | 2020.2 | 0.0 | 0.0 | 260.6 | 192.3 |
| 30 | 1579.0 | 9090.9 | 436.7 | 1041.7 | 3662.5 | 1551.7 | 30 | 0.0 | 2020.2 | 0.0 | 0.0 | 252.7 | 210.8 |
| | | | | | | | 31 | 0.0 | 2020.2 | 0.0 | 0.0 | 274.4 | 231.2 |

SARS-CoV-2 = severe acute respiratory syndrome coronavirus 2, P2 = period 2 (November 1, 2021 to December 31, 2021), Data are raw daily 7-day incidences extrapolated to 100,000 persons, Mo. = month, M.S. = middle school (students aged 13.1 ± 1.3 years old), H.S. = 4-year high school (students aged 16.9 ± 1.2 years old), AGES = Austrian Agency for Health and Food Safety, GB = students in school classes with a general school branch, SF = students in school classes with sport focus, I = 7 days incidence for SARS-CoV-2 infections per 100,000 inhabitants in Carinthia (AUT) for children aged 5 to 14 years (https://covid19-dashboard.ages.at/), II = 7 days incidence for SARS-CoV-2 infections per 100,000 inhabitants in Carinthia (AUT) for adolescent aged 15 to 24 years (https://covid19-dashboard.ages.at/), AUT = Austria, n = Study population, Nov.21 = November 2021, Dec.21 = December 2021.

**eTable 10.** Detailed 7-day incidences for detected SARS-CoV-2 infections for P3 (Jan.22 and Feb.22).

| Mo. | M.S. | | H.S. | | AGES | | Mo. | M.S. | | H.S. | | AGES | |
|---|---|---|---|---|---|---|---|---|---|---|---|---|---|
| Jan.22 | GB (n=190) | SF (n=99) | GB (n=229) | SF (n=96) | I | II | Feb.22 | GB (n=190) | SF (n=99) | GB (n=229) | SF (n=96) | I | II |
| 1 | 0.0 | 2020.2 | 436.7 | 1041.7 | 260.6 | 240.4 | 1 | 3684.2 | 11111.1 | 3930.1 | 7291.7 | 4282.4 | 3238.3 |
| 2 | 0.0 | 1010.1 | 436.7 | 1041.7 | 260.6 | 227.5 | 2 | 4210.5 | 11111.1 | 4366.8 | 8333.3 | 4207.4 | 3258.7 |
| 3 | 0.0 | 1010.1 | 436.7 | 2083.3 | 238.9 | 257.1 | 3 | 4736.8 | 8080.8 | 3930.1 | 6250.0 | 4262.7 | 3184.7 |
| 4 | 0.0 | 1010.1 | 436.7 | 2083.3 | 258.6 | 369.9 | 4 | 5263.2 | 8080.8 | 3930.1 | 7291.7 | 4641.8 | 3454.7 |
| 5 | 526.3 | 0.0 | 873.4 | 2083.3 | 292.2 | 519.7 | 5 | 5789.5 | 9090.9 | 4366.8 | 7291.7 | 4866.8 | 3711.8 |
| 6 | 526.3 | 0.0 | 1310.0 | 2083.3 | 321.8 | 654.7 | 6 | 5789.5 | 10101.0 | 3056.8 | 8333.3 | 5074.1 | 3904.1 |
| 7 | 526.3 | 1010.1 | 1746.7 | 2083.3 | 347.5 | 712.0 | 7 | 5263.2 | 8080.8 | 3056.8 | 5208.3 | 5127.4 | 3867.1 |
| 8 | 526.3 | 1010.1 | 1746.7 | 2083.3 | 408.7 | 865.5 | 8 | 4210.5 | 11111.1 | 2620.1 | 5208.3 | 5528.2 | 4218.5 |
| 9 | 1052.6 | 2020.2 | 2183.4 | 2083.3 | 456.1 | 950.6 | 9 | 4736.8 | 8080.8 | 3493.5 | 4166.7 | 5873.8 | 4446.0 |
| 10 | 3157.9 | 3030.3 | 2183.4 | 2083.3 | 586.4 | 1109.7 | 10 | 4210.5 | 11111.1 | 3930.1 | 5208.3 | 5794.8 | 4532.9 |
| 11 | 4210.5 | 4040.4 | 2183.4 | 3125.0 | 758.2 | 1213.2 | 11 | 3157.9 | 11111.1 | 3930.1 | 4166.7 | 5348.6 | 4270.3 |
| 12 | 3684.2 | 4040.4 | 1746.7 | 4166.7 | 807.5 | 1248.4 | 12 | 3684.2 | 10101.0 | 3056.8 | 4166.7 | 4924.1 | 4016.9 |
| 13 | 4210.5 | 4040.4 | 1310.0 | 5208.3 | 935.9 | 1294.6 | 13 | 4736.8 | 9090.9 | 3493.5 | 2083.3 | 4714.8 | 3885.6 |
| 14 | 4210.5 | 3030.3 | 873.4 | 5208.3 | 1028.6 | 1388.9 | 14 | 5263.2 | 8080.8 | 3493.5 | 2083.3 | 4507.5 | 3918.9 |
| 15 | 4210.5 | 3030.3 | 1746.7 | 6250.0 | 1129.3 | 1416.7 | 15 | 5263.2 | 6060.6 | 5240.2 | 1041.7 | 3753.3 | 3476.9 |
| 16 | 3684.2 | 2020.2 | 1310.0 | 10416.7 | 1239.9 | 1470.3 | 16 | 4210.5 | 5050.5 | 3930.1 | 1041.7 | 3016.8 | 3208.7 |
| 17 | 1579.0 | 2020.2 | 2620.1 | 10416.7 | 1342.6 | 1477.7 | 17 | 4736.8 | 2020.2 | 3493.5 | 0.0 | 2521.3 | 3001.6 |
| 18 | 1052.6 | 2020.2 | 3056.8 | 11458.3 | 1504.5 | 1631.2 | 18 | 5263.2 | 2020.2 | 4366.8 | 0.0 | 2270.5 | 2890.6 |
| 19 | 1579.0 | 2020.2 | 4366.8 | 13541.7 | 1836.2 | 1768.0 | 19 | 5789.5 | 2020.2 | 4366.8 | 1041.7 | 2246.8 | 2850.0 |
| 20 | 1052.6 | 3030.3 | 5240.2 | 14583.3 | 2071.1 | 1919.7 | 20 | 6842.1 | 2020.2 | 4366.8 | 1041.7 | 2260.7 | 2781.5 |
| 21 | 1579.0 | 5050.5 | 5676.9 | 18750.0 | 2631.8 | 2243.3 | 21 | 7368.4 | 3030.3 | 4366.8 | 1041.7 | 2321.9 | 2740.8 |
| 22 | 2105.3 | 6060.6 | 4366.8 | 19791.7 | 2825.3 | 2346.9 | 22 | 7894.7 | 2020.2 | 3493.5 | 1041.7 | 2689.1 | 2905.4 |
| 23 | 2631.6 | 7070.7 | 4366.8 | 16666.7 | 2983.3 | 2485.6 | 23 | 7894.7 | 3030.3 | 4803.5 | 1041.7 | 2935.9 | 2905.4 |
| 24 | 2631.6 | 7070.7 | 3056.8 | 15625.0 | 3082.0 | 2544.8 | 24 | 6315.8 | 3030.3 | 5676.9 | 1041.7 | 2977.4 | 2944.3 |
| 25 | 2631.6 | 6060.6 | 3056.8 | 14583.3 | 3482.8 | 2698.3 | 25 | 6842.1 | 2020.2 | 5240.2 | 1041.7 | 3208.4 | 3084.8 |
| 26 | 2105.3 | 10101.0 | 1746.7 | 11458.3 | 3816.5 | 2872.1 | 26 | 5263.2 | 3030.3 | 5240.2 | 0.0 | 3172.8 | 2977.6 |
| 27 | 3157.9 | 12121.2 | 1310.0 | 11458.3 | 4280.4 | 3158.8 | 27 | 3157.9 | 3030.3 | 4803.5 | 0.0 | 3149.1 | 2938.7 |
| 28 | 3157.9 | 11111.1 | 873.4 | 7291.7 | 4185.7 | 3105.2 | 28 | 2631.6 | 2020.2 | 4366.8 | 0.0 | 3101.7 | 2931.3 |
| 29 | 2631.6 | 10101.0 | 1310.0 | 4166.7 | 4302.2 | 3171.8 | | | | | | | |
| 30 | 2105.3 | 9090.9 | 2620.1 | 4166.7 | 4239.0 | 3155.1 | | | | | | | |
| 31 | 2631.6 | 11111.1 | 3493.5 | 7291.7 | 4339.7 | 3260.5 | | | | | | | |

SARS-CoV-2 = severe acute respiratory syndrome coronavirus 2, , P3 = period 3 (January 1, 2022 to February 28, 2022), Data are raw daily 7-day incidences extrapolated to 100,000 persons, Mo. = month, M.S. = middle school (students aged 13.1 ± 1.3 years old), H.S. = 4-year high school (students aged 16.9 ± 1.2 years old), AGES = Austrian Agency for Health and Food Safety, GB = students in school classes with a general school branch, SF = students in school classes with sport focus, I = 7 days incidence for SARS-CoV-2 infections per 100,000 inhabitants in Carinthia (AUT) for children aged 5 to 14 years (https://covid19-dashboard.ages.at/), II = 7 days incidence for SARS-CoV-2 infections per 100,000 inhabitants in Carinthia (AUT) for adolescent aged 15 to 24 years (https://covid19-dashboard.ages.at/), AUT = Austria, n = Study population, Jan.22 = January 2022, Feb.22 = February 2022.

**eTable 11.** Detailed 7-day incidences for detected SARS-CoV-2 infections for P4 (Mar.22 and Apr.22).

| Mo. | M.S. | | H.S. | | AGES | | Mo. | M.S. | | H.S. | | AGES | |
|---|---|---|---|---|---|---|---|---|---|---|---|---|---|
| Mar.22 | GB (n=190) | SF (n=99) | GB (n=229) | SF (n=96) | I | II | Apr.22 | GB (n=190) | SF (n=99) | GB (n=229) | SF (n=96) | I | II |
| 1 | 1579.0 | 3030.3 | 4803.5 | 0.0 | 3018.8 | 2877.7 | 1 | 1052.6 | 2020.2 | 1746.7 | 1041.7 | 1891.4 | 1686.7 |
| 2 | 2631.6 | 2020.2 | 3930.1 | 0.0 | 3403.8 | 3001.6 | 2 | 1052.6 | 2020.2 | 1746.7 | 1041.7 | 1842.1 | 1579.4 |
| 3 | 3157.9 | 2020.2 | 3056.8 | 0.0 | 3474.9 | 3058.9 | 3 | 2631.6 | 2020.2 | 1746.7 | 1041.7 | 1776.9 | 1475.8 |
| 4 | 2105.3 | 3030.3 | 3056.8 | 1041.7 | 3747.4 | 3138.5 | 4 | 2105.3 | 3030.3 | 873.4 | 1041.7 | 1660.4 | 1351.9 |
| 5 | 2105.3 | 2020.2 | 3930.1 | 1041.7 | 3861.9 | 3279.0 | 5 | 2105.3 | 2020.2 | 873.4 | 2083.3 | 1563.7 | 1270.6 |
| 6 | 2105.3 | 2020.2 | 3930.1 | 1041.7 | 3913.2 | 3297.5 | 6 | 2105.3 | 2020.2 | 1310.0 | 1041.7 | 1350.5 | 1091.2 |
| 7 | 1579.0 | 2020.2 | 3930.1 | 1041.7 | 4102.7 | 3373.3 | 7 | 2105.3 | 2020.2 | 1746.7 | 1041.7 | 1283.3 | 1000.5 |
| 8 | 2105.3 | 1010.1 | 2183.4 | 1041.7 | 4539.1 | 3669.2 | 8 | 2105.3 | 1010.1 | 1310.0 | 1041.7 | 1082.0 | 882.2 |
| 9 | 3157.9 | 1010.1 | 3056.8 | 1041.7 | 4477.9 | 3978.1 | 9 | 1579.0 | 1010.1 | 1310.0 | 1041.7 | 1056.3 | 869.2 |
| 10 | 3684.2 | 1010.1 | 4366.8 | 1041.7 | 4612.1 | 4266.6 | 10 | 526.3 | 1010.1 | 1310.0 | 1041.7 | 1024.7 | 832.2 |
| 11 | 5263.2 | 0.0 | 3930.1 | 0.0 | 4675.3 | 4488.5 | 11 | 526.3 | 0.0 | 1310.0 | 1041.7 | 981.3 | 795.3 |
| 12 | 6842.1 | 0.0 | 3056.8 | 0.0 | 4734.5 | 4625.4 | 12 | 526.3 | 0.0 | 1310.0 | 0.0 | 675.2 | 630.7 |
| 13 | 6842.1 | 0.0 | 3056.8 | 0.0 | 4805.6 | 4725.3 | 13 | 526.3 | 0.0 | 873.4 | 0.0 | 519.3 | 536.3 |
| 14 | 8421.1 | 0.0 | 2620.1 | 0.0 | 4740.5 | 4778.9 | 14 | 526.3 | 0.0 | 873.4 | 0.0 | 493.6 | 499.3 |
| 15 | 8947.4 | 0.0 | 3493.5 | 0.0 | 4647.7 | 4817.7 | 15 | 1052.6 | 0.0 | 436.7 | 0.0 | 351.4 | 427.2 |
| 16 | 8947.4 | 0.0 | 4366.8 | 0.0 | 4849.1 | 4814.0 | 16 | 1052.6 | 0.0 | 436.7 | 0.0 | 333.7 | 386.5 |
| 17 | 8421.1 | 0.0 | 3493.5 | 0.0 | 4701.0 | 4630.9 | 17 | 526.3 | 0.0 | 436.7 | 1041.7 | 319.8 | 368.0 |
| 18 | 7368.4 | 0.0 | 4366.8 | 0.0 | 4651.6 | 4514.4 | 18 | 526.3 | 0.0 | 436.7 | 1041.7 | 315.9 | 364.3 |
| 19 | 5789.5 | 0.0 | 4366.8 | 0.0 | 4556.9 | 4396.1 | 19 | 526.3 | 0.0 | 873.4 | 1041.7 | 331.7 | 331.0 |
| 20 | 5789.5 | 0.0 | 6113.5 | 0.0 | 4432.5 | 4360.9 | 20 | 526.3 | 0.0 | 1310.0 | 1041.7 | 475.8 | 408.7 |
| 21 | 4736.8 | 1010.1 | 7423.6 | 0.0 | 4306.1 | 4296.2 | 21 | 526.3 | 0.0 | 873.4 | 1041.7 | 467.9 | 412.4 |
| 22 | 4210.5 | 1010.1 | 7860.3 | 0.0 | 4183.7 | 4081.7 | 22 | 0.0 | 0.0 | 873.4 | 1041.7 | 489.6 | 432.8 |
| 23 | 2105.3 | 2020.2 | 6550.2 | 0.0 | 3761.2 | 3650.8 | 23 | 0.0 | 0.0 | 873.4 | 2083.3 | 483.7 | 421.7 |
| 24 | 2105.3 | 2020.2 | 6113.5 | 0.0 | 3696.0 | 3382.6 | 24 | 0.0 | 0.0 | 873.4 | 1041.7 | 481.7 | 432.8 |
| 25 | 1579.0 | 2020.2 | 5240.2 | 0.0 | 3194.5 | 3040.4 | 25 | 0.0 | 0.0 | 1310.0 | 1041.7 | 501.5 | 427.2 |
| 26 | 2105.3 | 2020.2 | 5240.2 | 0.0 | 3034.6 | 2861.1 | 26 | 0.0 | 0.0 | 873.4 | 1041.7 | 517.3 | 440.2 |
| 27 | 2105.3 | 2020.2 | 3493.5 | 0.0 | 2890.5 | 2718.6 | 27 | 0.0 | 0.0 | 436.7 | 1041.7 | 404.7 | 356.9 |
| 28 | 2105.3 | 1010.1 | 3056.8 | 0.0 | 2787.8 | 2537.4 | 28 | 0.0 | 0.0 | 436.7 | 1041.7 | 430.4 | 351.4 |
| 29 | 1579.0 | 2020.2 | 1746.7 | 0.0 | 2351.5 | 2171.2 | 29 | 0.0 | 0.0 | 436.7 | 1041.7 | 424.5 | 318.1 |
| 30 | 1579.0 | 1010.1 | 873.4 | 1041.7 | 2100.7 | 1997.4 | 30 | 0.0 | 1010.1 | 436.7 | 0.0 | 446.2 | 303.3 |
| 31 | 1052.6 | 1010.1 | 873.4 | 1041.7 | 1838.1 | 1784.7 | | | | | | | |

SARS-CoV-2 = severe acute respiratory syndrome coronavirus 2, , P4 = period 4 (March 1, 2022 to April 30, 2022), Data are raw daily 7-day incidences extrapolated to 100,000 persons, Mo. = month, M.S. = middle school (students aged 13.1 ± 1.3 years old), H.S. = 4-year high school (students aged 16.9 ± 1.2 years old), AGES = Austrian Agency for Health and Food Safety, GB = students in school classes with a general school branch, SF = students in school classes with sport focus, I = 7 days incidence for SARS-CoV-2 infections per 100,000 inhabitants in Carinthia (AUT) for children aged 5 to 14 years (https://covid19-dashboard.ages.at/), II = 7 days incidence for SARS-CoV-2 infections per 100,000 inhabitants in Carinthia (AUT) for adolescent aged 15 to 24 years (https://covid19-dashboard.ages.at/), AUT = Austria, n = Study population, Mar.22 = March 2022, Apr.22 = April 2022.

**eTable 12:** Potential SARS-CoV-2 infection in school classes GB vs. SF classes, using hypothesized generation time - 2 days.

| Time Period | school grade | Category | HTPPI = GT 2D | | | | | | |
|---|---|---|---|---|---|---|---|---|---|
| | | | PI, No. | | $X^2$ | P Value [b] | p-lvl | φ | OR (95%CI) |
| | | | yes | no | | | | | |
| P1 | All (n=614) | GB (n=419) | 0 | 419 | INC | | | | |
| | | SF (n=185) | 0 | 195 | | | | | |
| | M.S. (n=289) | GB (n=190) | 0 | 190 | INC | | | | |
| | | SF (n=99) | 0 | 99 | | | | | |
| | H.S. (n=325) | GB (n=229) | 0 | 229 | INC | | | | |
| | | SF (n=96) | 0 | 96 | | | | | |
| P2 | All (n=614) | GB (n=419) | 3 | 416 | 6.859 [a] | 0.014 | * | 0.11 | 5.16 (1.32 to 20.19) |
| | | SF (n=185) | 7 | 188 | | | | | |
| | M.S. (n=289) | GB (n=190) | 0 | 190 | INC | | | | |
| | | SF (n=99) | 5 | 94 | | | | | |
| | H.S. (n=325) | GB (n=229) | 3 | 226 | 0.267 [a] | 0.63 | | 0.03 | 1.60 (0.26 to 9.75) |
| | | SF (n=96) | 2 | 94 | | | | | |
| P3 | All (n=614) | GB (n=419) | 26 | 393 | 30.066 | <.001 | *** | 0.22 | 4.02 (2.38 to 6.81) |
| | | SF (n=185) | 41 | 154 | | | | | |
| | M.S. (n=289) | GB (n=190) | 12 | 178 | 15.864 | <.001 | *** | 0.23 | 4.24 (2.00 to 8.99) |
| | | SF (n=99) | 22 | 77 | | | | | |
| | H.S. (n=325) | GB (n=229) | 14 | 215 | 13.872 | <.001 | *** | 0.21 | 3.79 (1.81 to 7.93) |
| | | SF (n=96) | 19 | 77 | | | | | |
| P4 | All (n=614) | GB (n=419) | 28 | 391 | INC | | | | |
| | | SF (n=185) | 0 | 195 | | | | | |
| | M.S. (n=289) | GB (n=190) | 14 | 176 | INC | | | | |
| | | SF (n=99) | 0 | 99 | | | | | |
| | H.S. (n=325) | GB (n=229) | 14 | 215 | INC | | | | |
| | | SF (n=96) | 0 | 96 | | | | | |

a = 1 cell (25%) or 2 cells (50,0%) have expected count less than 5; b = If ( a ) occurred, Fisher's Exact Test was used.
SARS-CoV-2 = severe acute respiratory syndrome coronavirus 2, HTPPI = hypothetical time period assumed for potential infections, GT = generation time, D = days, PI = potential infection, No = absolute Number of children, $X^2$ = Chi-Square Test value, φ = Effect size Phi , p-lvl (P Value level) * = P < 0.05, ** = P < 0.01, *** = P < 0.001, p-lvl = significance level, OR = Odds ratio for potential SARS-CoV-2 infection in school classes with sport focus vs. school classes without sport focus, using different hypothesized generation time (with 1 for students in classes without sport focus), CI = Confidence Interval, P1 = period 1 (September 13, 2021 to October 31, 2021) P2 = period 2 (November 1, 2021 to December 31, 2021), P3 = period 3 (January 1, 2022 to February 28, 2022), P4 = period 4 (March 1, 2022 to April 30, 2022), M.S. = middle school (students aged 13.1 ± 1.3 years old), H.S. = 4-year high school (students aged 16.9 ± 1.2 years old), GB = students in school classes with a general school branch, SF = students in school classes with sport focus, n = Study population, INC = insufficient number of potential infection cases.

**eTable 13:** Potential SARS-CoV-2 infection in school classes GB vs. SF classes, using hypothesized generation time - 4 days.

| Time Period | school grade | Category | HTPPI = GT 4D | | | | | | |
|---|---|---|---|---|---|---|---|---|---|
| | | | PI, No. | | $X^2$ | P Value [b] | p-lvl | φ | OR (95%CI) |
| | | | yes | no | | | | | |
| P1 | All (n=614) | GB (n=419) | 0 | 419 | INC | | | | |
| | | SF (n=185) | 0 | 195 | | | | | |
| | M.S. (n=289) | GB (n=190) | 0 | 190 | INC | | | | |
| | | SF (n=99) | 0 | 99 | | | | | |
| | H.S. (n=325) | GB (n=229) | 0 | 229 | INC | | | | |
| | | SF (n=96) | 0 | 96 | | | | | |
| P2 | All (n=614) | GB (n=419) | 8 | 411 | 4.845 | 0.028 | * | 0.09 | 2.78 (1.08 to 7.15) |
| | | SF (n=185) | 10 | 185 | | | | | |
| | M.S. (n=289) | GB (n=190) | 1 | 189 | 8.435 [a] | 0.007 | ** | 0.17 | 12.19 (1.45 to 102.76) |
| | | SF (n=99) | 6 | 93 | | | | | |
| | H.S. (n=325) | GB (n=229) | 7 | 222 | 0.255 [a] | 0.74 | | 0.03 | 1.38 (0.39 to 4.82) |
| | | SF (n=96) | 4 | 92 | | | | | |
| P3 | All (n=614) | GB (n=419) | 39 | 380 | 40.042 | <.001 | *** | 0.26 | 4.03 (2.56 to 6.31) |
| | | SF (n=185) | 57 | 138 | | | | | |
| | M.S. (n=289) | GB (n=190) | 18 | 172 | 17.205 | <.001 | *** | 0.24 | 3.77 (1.96 to 7.24) |
| | | SF (n=99) | 28 | 71 | | | | | |
| | H.S. (n=325) | GB (n=229) | 21 | 208 | 22.998 | <.001 | *** | 0.27 | 4.29 (2.29 to 8.01) |
| | | SF (n=96) | 29 | 67 | | | | | |
| P4 | All (n=614) | GB (n=419) | 37 | 382 | 15.857 | <.001 | *** | -0.16 | 0.05 (0.01 to 0.39) |
| | | SF (n=185) | 1 | 194 | | | | | |
| | M.S. (n=289) | GB (n=190) | 17 | 173 | 7.021 | 0.008 | ** | -0.16 | 0.10 (0.01 to 0.79) |
| | | SF (n=99) | 1 | 98 | | | | | |
| | H.S. (n=325) | GB (n=229) | 20 | 209 | 8.934 | 0.003 | ** | -0.16 | 0.10 (0.01 to 0.79) |
| | | SF (n=96) | 0 | 96 | | | | | |

a = 1 cell (25%) or 2 cells (50,0%) have expected count less than 5; b = If ( a ) occurred, Fisher's Exact Test was used.
SARS-CoV-2 = severe acute respiratory syndrome coronavirus 2, HTPPI = hypothetical time period assumed for potential infections, GT = generation time, D = days, PI = potential infection, No = absolute Number of children, $X^2$ = Chi-Square Test value, φ = Effect size Phi , p-lvl (P Value level) * = P < 0.05, ** = P < 0.01, *** = P < 0.001, p-lvl = significance level, OR = Odds ratio for potential SARS-CoV-2 infection in school classes with sport focus vs. school classes without sport focus, using different hypothesized generation time (with 1 for students in classes without sport focus), CI = Confidence Interval, P1 = period 1 (September 13, 2021 to October 31, 2021) P2 = period 2 (November 1, 2021 to December 31, 2021), P3 = period 3 (January 1, 2022 to February 28, 2022), P4 = period 4 (March 1, 2022 to April 30, 2022), M.S. = middle school (students aged 13.1 ± 1.3 years old), H.S. = 4-year high school (students aged 16.9 ± 1.2 years old), GB = students in school classes with a general school branch, SF = students in school classes with sport focus, n = Study population, INC = insufficient number of potential infection cases.

**eTable 14:** Potential SARS-CoV-2 infection in school classes GB vs. SF classes, using hypothesized generation time - 6 days.

| Time Period | school grade | Category | HTPPI = GT 6D | | | | | | |
|---|---|---|---|---|---|---|---|---|---|
| | | | PI, No. | | $X^2$ | P Value [b] | p-lvl | φ | OR (95%CI) |
| | | | yes | no | | | | | |
| P1 | All (n=614) | GB (n=419) | 0 | 419 | INC | | | | |
| | | SF (n=185) | 0 | 195 | | | | | |
| | M.S. (n=289) | GB (n=190) | 0 | 190 | INC | | | | |
| | | SF (n=99) | 0 | 99 | | | | | |
| | H.S. (n=325) | GB (n=229) | 0 | 229 | INC | | | | |
| | | SF (n=96) | 0 | 96 | | | | | |
| P2 | All (n=614) | GB (n=419) | 10 | 409 | 6.761 | 0.009 | ** | 0.11 | 2.92 (1.26 to 6.79) |
| | | SF (n=185) | 13 | 182 | | | | | |
| | M.S. (n=289) | GB (n=190) | 3 | 187 | 4.333 [a] | 0.07 | | 0.04 | 4.02 (0.98 to 16.44) |
| | | SF (n=99) | 6 | 93 | | | | | |
| | H.S. (n=325) | GB (n=229) | 7 | 222 | 2.943 [a] | 0.13 | | 0.09 | 2.49 (0.85 to 7.32) |
| | | SF (n=96) | 7 | 89 | | | | | |
| P3 | All (n=614) | GB (n=419) | 45 | 374 | 39.313 | <.001 | *** | 0.25 | 3.78 (2.45 to 5.83) |
| | | SF (n=185) | 61 | 134 | | | | | |
| | M.S. (n=289) | GB (n=190) | 21 | 169 | 16.597 | <.001 | *** | 0.24 | 3.50 (1.88 to 6.53) |
| | | SF (n=99) | 30 | 69 | | | | | |
| | H.S. (n=325) | GB (n=229) | 24 | 205 | 22.889 | <.001 | *** | 0.27 | 4.08 (2.23 to 7.43) |
| | | SF (n=96) | 31 | 65 | | | | | |
| P4 | All (n=614) | GB (n=419) | 51 | 368 | 18.758 | <.001 | *** | -0.175 | 0.11 (0.04 to 0.37) |
| | | SF (n=185) | 3 | 192 | | | | | |
| | M.S. (n=289) | GB (n=190) | 23 | 167 | 8.377 | 0.004 | ** | -0.17 | 0.15 (0.04 to 0.65) |
| | | SF (n=99) | 2 | 97 | | | | | |
| | H.S. (n=325) | GB (n=229) | 28 | 201 | 10.414 | 0.001 | ** | -0.18 | 0.08 (0.01 to 0.56) |
| | | SF (n=96) | 1 | 95 | | | | | |

a = 1 cell (25%) or 2 cells (50,0%) have expected count less than 5; b = If ( a ) occurred, Fisher's Exact Test was used.
SARS-CoV-2 = severe acute respiratory syndrome coronavirus 2, HTPPI = hypothetical time period assumed for potential infections, GT = generation time, D = days, PI = potential infection, No = absolute Number of children, $X^2$ = Chi-Square Test value, φ = Effect size Phi , p-lvl (P Value level) * = P < 0.05, ** = P < 0.01, *** = P < 0.001, p-lvl = significance level, OR = Odds ratio for potential SARS-CoV-2 infection in school classes with sport focus vs. school classes without sport focus, using different hypothesized generation time (with 1 for students in classes without sport focus), CI = Confidence Interval, P1 = period 1 (September 13, 2021 to October 31, 2021) P2 = period 2 (November 1, 2021 to December 31, 2021), P3 = period 3 (January 1, 2022 to February 28, 2022), P4 = period 4 (March 1, 2022 to April 30, 2022), M.S. = middle school (students aged 13.1 ± 1.3 years old), H.S. = 4-year high school (students aged 16.9 ± 1.2 years old), GB = students in school classes with a general school branch, SF = students in school classes with sport focus, n = Study population, INC = insufficient number of potential infection cases.

**eTable 15:** Potential SARS-CoV-2 infection in school classes GB vs. SF classes, using hypothesized generation time - 8 days.

| Time Period | school grade | Category | HTPPI = GT 8D | | | | | | |
|---|---|---|---|---|---|---|---|---|---|
| | | | PI, No. | | $X^2$ | P Value [b] | p-lvl | φ | OR (95%CI) |
| | | | yes | no | | | | | |
| P1 | All (n=614) | GB (n=419) | 2 | 417 | INC | | | | |
| | | SF (n=185) | 0 | 195 | | | | | |
| | M.S. (n=289) | GB (n=190) | 0 | 190 | INC | | | | |
| | | SF (n=99) | 0 | 99 | | | | | |
| | H.S. (n=325) | GB (n=229) | 2 | 227 | INC | | | | |
| | | SF (n=96) | 0 | 96 | | | | | |
| P2 | All (n=614) | GB (n=419) | 11 | 408 | 7.066 | 0.008 | ** | 0.11 | 2.87 (1.28 to 6.44) |
| | | SF (n=185) | 14 | 181 | | | | | |
| | M.S. (n=289) | GB (n=190) | 3 | 187 | 5.876 [a] | 0.035 | * | 0.14 | 4.74 (1.20 to 18.76) |
| | | SF (n=99) | 7 | 92 | | | | | |
| | H.S. (n=325) | GB (n=229) | 8 | 221 | 2.217 [a] | 0.15 | | 0.08 | 2.17 (0.77 to 6.17) |
| | | SF (n=96) | 7 | 89 | | | | | |
| P3 | All (n=614) | GB (n=419) | 54 | 365 | 34.055 | <.001 | *** | 0.24 | 3.30 (2.18 to 4.99) |
| | | SF (n=185) | 64 | 131 | | | | | |
| | M.S. (n=289) | GB (n=190) | 24 | 166 | 13.377 | <.001 | *** | 0.22 | 3.01 (1.64 to 5.51) |
| | | SF (n=99) | 30 | 69 | | | | | |
| | H.S. (n=325) | GB (n=229) | 30 | 199 | 21.302 | <.001 | *** | 0.26 | 3.64 (2.06 to 6.42) |
| | | SF (n=96) | 34 | 62 | | | | | |
| P4 | All (n=614) | GB (n=419) | 56 | 363 | 19.319 | <.001 | *** | -0.18 | 0.14 (0.05 to 0.38) |
| | | SF (n=185) | 4 | 191 | | | | | |
| | M.S. (n=289) | GB (n=190) | 25 | 165 | 7.630 | 0.006 | ** | -0.16 | 0.21 (0.06 to 0.70) |
| | | SF (n=99) | 3 | 96 | | | | | |
| | H.S. (n=325) | GB (n=229) | 31 | 198 | 11.898 | 0.001 | ** | -0.19 | 0.07 (0.01 to 0.50) |
| | | SF (n=96) | 1 | 95 | | | | | |

a = 1 cell (25%) or 2 cells (50,0%) have expected count less than 5; b = If ( a ) occurred, Fisher's Exact Test was used.
SARS-CoV-2 = severe acute respiratory syndrome coronavirus 2, HTPPI = hypothetical time period assumed for potential infections, GT = generation time, D = days, PI = potential infection, No = absolute Number of children, $X^2$ = Chi-Square Test value, φ = Effect size Phi , p-lvl (P Value level) * = P < 0.05, ** = P < 0.01, *** = P < 0.001, p-lvl = significance level, OR = Odds ratio for potential SARS-CoV-2 infection in school classes with sport focus vs. school classes without sport focus, using different hypothesized generation time (with 1 for students in classes without sport focus), CI = Confidence Interval, P1 = period 1 (September 13, 2021 to October 31, 2021) P2 = period 2 (November 1, 2021 to December 31, 2021), P3 = period 3 (January 1, 2022 to February 28, 2022), P4 = period 4 (March 1, 2022 to April 30, 2022), M.S. = middle school (students aged 13.1 ± 1.3 years old), H.S. = 4-year high school (students aged 16.9 ± 1.2 years old), GB = students in school classes with a general school branch, SF = students in school classes with sport focus, n = Study population, INC = insufficient number of potential infection cases.

**eTable 16.** Binary logistic regression for Potential SARS-Cov-2 contagions in the classroom and generation times 4 and 6, for subgroup sex .

| Time Period | school grade | Category | Sex | HTPPI = GT 4D | | | HTPPI = GT 6D | | |
|---|---|---|---|---|---|---|---|---|---|
| | | | | PI | | OR (95%CI) | PI | | OR (95%CI) |
| | | | | yes | no | | yes | no | |
| P1 | All (n=614) | ♂ (n=349) | A (n=203) | 0 | 203 | INC | 0 | 203 | INC |
| | | | B (n=146) | 0 | 146 | | 0 | 146 | |
| | | ♀ (n=265) | A (n=216) | 0 | 216 | INC | 0 | 216 | INC |
| | | | B (n=49) | 0 | 49 | | 0 | 49 | |
| | M.S. (n=289) | ♂ (n=178) | A (n=106) | 0 | 106 | INC | 0 | 106 | INC |
| | | | B (n=72) | 0 | 72 | | 0 | 72 | |
| | | ♀ (n=111) | A (n=84) | 0 | 84 | INC | 0 | 84 | INC |
| | | | B (n=27) | 0 | 27 | | 0 | 27 | |
| | H.S. (n=325) | ♂ (n=171) | A (n=97) | 0 | 97 | INC | 0 | 97 | INC |
| | | | B (n=74) | 0 | 74 | | 0 | 74 | |
| | | ♀ (n=154) | A (n=132) | 0 | 132 | INC | 0 | 132 | INC |
| | | | B (n=22) | 0 | 22 | | 0 | 22 | |
| P2 | All (n=614) | ♂ (n=349) | A (n=203) | 4 | 199 | 3.66 (1.12 to 11.90) | 5 | 198 | 3.22 (1.10 to 9.50) |
| | | | B (n=146) | 10 | 136 | | 11 | 135 | |
| | | ♀ (n=265) | A (n=216) | 4 | 212 | 0.81 (0.77 to 0.86) | 5 | 211 | 1.15 (0.34 to 9.54) |
| | | | B (n=49) | 0 | 49 | | 2 | 47 | |
| | M.S. (n=289) | ♂ (n=178) | A (n=106) | 1 | 105 | 9.55 (1.12 to 81.07) | 2 | 104 | 4.73 (0.93 to 24.12) |
| | | | B (n=72) | 6 | 66 | | 6 | 66 | |
| | | ♀ (n=111) | A (n=84) | 0 | 84 | INC | 1 | 83 | INC |
| | | | B (n=27) | 0 | 27 | | 0 | 27 | |
| | H.S. (n=325) | ♂ (n=171) | A (n=97) | 3 | 94 | 1.79 (0.39 to 8.26) | 3 | 94 | 2.27 (0.53 to 9.82) |
| | | | B (n=74) | 4 | 70 | | 5 | 69 | |
| | | ♀ (n=154) | A (n=132) | 4 | 128 | 0.85 (0.80 to 0.91) | 4 | 128 | 3.20 (0.55 to 18.63) |
| | | | B (n=22) | 0 | 22 | | 2 | 20 | |
| P3 | All (n=614) | ♂ (n=349) | A (n=203) | 14 | 189 | 4.58 (2.37 to 8.85) | 18 | 185 | 3.88 (2.12 to 7.11) |
| | | | B (n=146) | 37 | 109 | | 40 | 106 | |
| | | ♀ (n=265) | A (n=216) | 25 | 191 | 5.27 (2.60 to 10.67) | 27 | 189 | 5.25 (2.62 to 10.52) |
| | | | B (n=49) | 20 | 29 | | 21 | 28 | |
| | M.S. (n=289) | ♂ (n=178) | A (n=106) | 8 | 98 | 3.50 (1.41 to 8.70) | 10 | 96 | 2.97 (1.27 to 6.93) |
| | | | B (n=72) | 16 | 56 | | 17 | 55 | |
| | | ♀ (n=111) | A (n=84) | 10 | 74 | 5.92 (2.17 to 16.19) | 11 | 73 | 6.16 (2.30 to 16.51) |
| | | | B (n=27) | 12 | 15 | | 13 | 14 | |
| | H.S. (n=325) | ♂ (n=171) | A (n=97) | 6 | 91 | 6.01 (2.28 to 15.83) | 8 | 89 | 5.02 (2.09 to 12.04) |
| | | | B (n=74) | 21 | 53 | | 23 | 51 | |
| | | ♀ (n=154) | A (n=132) | 15 | 117 | 4.46 (1.61 to 12.38) | 16 | 116 | 4.14 (1.50 to 11.42) |
| | | | B (n=22) | 8 | 14 | | 8 | 14 | |
| P4 | All (n=614) | ♂ (n=349) | A (n=203) | 14 | 189 | 0.56 (0.51 to 0.62) | 21 | 182 | 0.12 (0.03 to 0.52) |
| | | | B (n=146) | 0 | 146 | | 2 | 144 | |
| | | ♀ (n=265) | A (n=216) | 23 | 193 | 0.18 (0.02 to 1.33) | 30 | 186 | 0.13 (0.02 to 0.97) |
| | | | B (n=49) | 1 | 48 | | 1 | 48 | |
| | M.S. (n=289) | ♂ (n=178) | A (n=106) | 8 | 98 | 0.58 (0.51 to 0.66) | 11 | 95 | 0.12 (0.02 to 0.96) |
| | | | B (n=72) | 0 | 72 | | 1 | 71 | |
| | | ♀ (n=111) | A (n=84) | 9 | 75 | 0.32 (0.04 to 2.65) | 12 | 72 | 0.23 (0.03 to 1.86) |
| | | | B (n=27) | 1 | 26 | | 1 | 26 | |
| | H.S. (n=325) | ♂ (n=171) | A (n=97) | 6 | 91 | 0.55 (0.48 to 0.63) | 10 | 87 | 0.12 (0.02 to 0.95) |
| | | | B (n=74) | 0 | 74 | | 1 | 73 | |
| | | ♀ (n=154) | A (n=132) | 14 | 118 | INC | 14 | 118 | INC |
| | | | B (n=22) | 0 | 22 | | 0 | 22 | |

SARS-CoV-2 = severe acute respiratory syndrome coronavirus 2, HTPPI = hypothetical time period assumed for potential infections, GT = generation time, D = days, PI = potential infection, No = absolute Number of children, OR = Odds ratio for potential SARS-CoV-2 infection in school classes with sport focus vs. school classes without sport focus, using different hypothesized generation time (with 1 for students in classes without sport focus), CI = Confidence Interval, ♂ = Boys, ♀ = Girls, P1 = period 1 (September 13, 2021 to October 31, 2021) P2 = period 2 (November 1, 2021 to December 31, 2021), P3 = period 3 (January 1, 2022 to February 28, 2022), P4 = period 4 (March 1, 2022 to April 30, 2022), M.S. = middle school (students aged 13.1 ± 1.3 years old), H.S. = 4-year high school (students aged 16.9 ± 1.2 years old), GB = students in school classes with a general school branch, SF = students in school classes with sport focus, n = Study population, INC = insufficient number of potential infection cases.

**eTable 17.** Binary logistic regression for Potential SARS-Cov-2 contagions in the classroom and generation times 2 and 8, for subgroup sex .

| Time Period | school grade | Category | Sex | HTPPI = GT 2D | | | HTPPI = GT 8D | | |
|---|---|---|---|---|---|---|---|---|---|
| | | | | PI | | OR (95%CI) | PI | | OR (95%CI) |
| | | | | yes | no | | yes | no | |
| P1 | All (n=614) | ♂ (n=349) | A (n=203) | 0 | 203 | INC | 1 | 202 | INC |
| | | | B (n=146) | 0 | 146 | | 0 | 146 | |
| | | ♀ (n=265) | A (n=216) | 0 | 216 | INC | 1 | 215 | INC |
| | | | B (n=49) | 0 | 49 | | 0 | 49 | |
| | M.S. (n=289) | ♂ (n=178) | A (n=106) | 0 | 106 | INC | 0 | 106 | INC |
| | | | B (n=72) | 0 | 72 | | 0 | 72 | |
| | | ♀ (n=111) | A (n=84) | 0 | 84 | INC | 0 | 84 | INC |
| | | | B (n=27) | 0 | 27 | | 0 | 27 | |
| | H.S. (n=325) | ♂ (n=171) | A (n=97) | 0 | 97 | INC | 1 | 96 | INC |
| | | | B (n=74) | 0 | 74 | | 0 | 74 | |
| | | ♀ (n=154) | A (n=132) | 0 | 132 | INC | 1 | 131 | INC |
| | | | B (n=22) | 0 | 22 | | 0 | 22 | |
| P2 | All (n=614) | ♂ (n=349) | A (n=203) | 2 | 201 | 5.06 (1.04 to 24.73) | 5 | 198 | 3.55 (1.22 to 10.30) |
| | | | B (n=146) | 7 | 139 | | 12 | 135 | |
| | | ♀ (n=265) | A (n=216) | 1 | 215 | INC | 6 | 210 | 1.49 (0.30 to 7.61) |
| | | | B (n=49) | 0 | 49 | | 2 | 47 | |
| | M.S. (n=289) | ♂ (n=178) | A (n=106) | 0 | 106 | INC | 2 | 104 | 5.60 (1.13 to 27.79) |
| | | | B (n=72) | 5 | 67 | | 7 | 65 | |
| | | ♀ (n=111) | A (n=84) | 0 | 84 | INC | 1 | 83 | INC |
| | | | B (n=27) | 0 | 27 | | 0 | 27 | |
| | H.S. (n=325) | ♂ (n=171) | A (n=97) | 2 | 95 | 1.32 (0.18 to 9.59) | 3 | 94 | 2.27 (0.53 to 9.82) |
| | | | B (n=74) | 2 | 72 | | 5 | 69 | |
| | | ♀ (n=154) | A (n=132) | 1 | 131 | INC | 5 | 127 | 2.54 (0.46 to 13.99) |
| | | | B (n=22) | 0 | 22 | | 2 | 20 | |
| P3 | All (n=614) | ♂ (n=349) | A (n=203) | 9 | 194 | 4.45 (2.01 to 9.86) | 21 | 182 | 3.50 (1.97 to 6.23) |
| | | | B (n=146) | 25 | 121 | | 42 | 104 | |
| | | ♀ (n=265) | A (n=216) | 17 | 199 | 5.68 (2.61 to 12.33) | 33 | 183 | 4.52 (2.30 to 8.87) |
| | | | B (n=49) | 16 | 33 | | 22 | 27 | |
| | M.S. (n=289) | ♂ (n=178) | A (n=106) | 6 | 100 | 3.67 (1.33 to 10.18) | 11 | 95 | 2.67 (1.17 to 6.11) |
| | | | B (n=72) | 13 | 59 | | 17 | 55 | |
| | | ♀ (n=111) | A (n=84) | 6 | 78 | 6.50 (2.05 to 20.59) | 13 | 71 | 5.07 (1.94 to 13.23) |
| | | | B (n=27) | 9 | 18 | | 13 | 14 | |
| | H.S. (n=325) | ♂ (n=171) | A (n=97) | 3 | 94 | 6.07 (1.64 to 22.37) | 10 | 87 | 4.44 (1.97 to 10.00) |
| | | | B (n=74) | 12 | 62 | | 25 | 49 | |
| | | ♀ (n=154) | A (n=132) | 11 | 121 | 5.13 (1.73 to 15.25) | 20 | 112 | 3.88 (1.46 to 10.27) |
| | | | B (n=22) | 7 | 15 | | 9 | 13 | |
| P4 | All (n=614) | ♂ (n=349) | A (n=203) | 11 | 192 | INC | 25 | 178 | 0.15 (0.04 to 0.51) |
| | | | B (n=146) | 0 | 146 | | 3 | 143 | |
| | | ♀ (n=265) | A (n=216) | 17 | 199 | INC | 31 | 185 | 0.12 (0.02 to 0.93) |
| | | | B (n=49) | 0 | 49 | | 1 | 48 | |
| | M.S. (n=289) | ♂ (n=178) | A (n=106) | 6 | 100 | INC | 13 | 93 | 0.20 (0.05 to 0.94) |
| | | | B (n=72) | 0 | 72 | | 2 | 72 | |
| | | ♀ (n=111) | A (n=84) | 8 | 76 | INC | 12 | 72 | 0.23 (0.03 to 1.86) |
| | | | B (n=27) | 0 | 27 | | 1 | 26 | |
| | H.S. (n=325) | ♂ (n=171) | A (n=97) | 5 | 92 | INC | 12 | 85 | 0.10 (0.01 to 0.76) |
| | | | B (n=74) | 0 | 74 | | 1 | 73 | |
| | | ♀ (n=154) | A (n=132) | 9 | 123 | INC | 19 | 113 | INC |
| | | | B (n=22) | 0 | 22 | | 0 | 22 | |

SARS-CoV-2 = severe acute respiratory syndrome coronavirus 2, HTPPI = hypothetical time period assumed for potential infections, GT = generation time, D = days, PI = potential infection, No = absolute Number of children, OR = Odds ratio for potential SARS-CoV-2 infection in school classes with sport focus vs. school classes without sport focus, using different hypothesized generation time (with 1 for students in classes without sport focus), CI = Confidence Interval, ♂ = Boys, ♀ = Girls, P1 = period 1 (September 13, 2021 to October 31, 2021) P2 = period 2 (November 1, 2021 to December 31, 2021), P3 = period 3 (January 1, 2022 to February 28, 2022), P4 = period 4 (March 1, 2022 to April 30, 2022), M.S. = middle school (students aged 13.1 ± 1.3 years old), H.S. = 4-year high school (students aged 16.9 ± 1.2 years old), GB = students in school classes with a general school branch, SF = students in school classes with sport focus, n = Study population, INC = insufficient number of potential infection cases.

**eFigure 1.** Cumulative percentage of children with SARS-CoV-2 infections, for subgroup sex

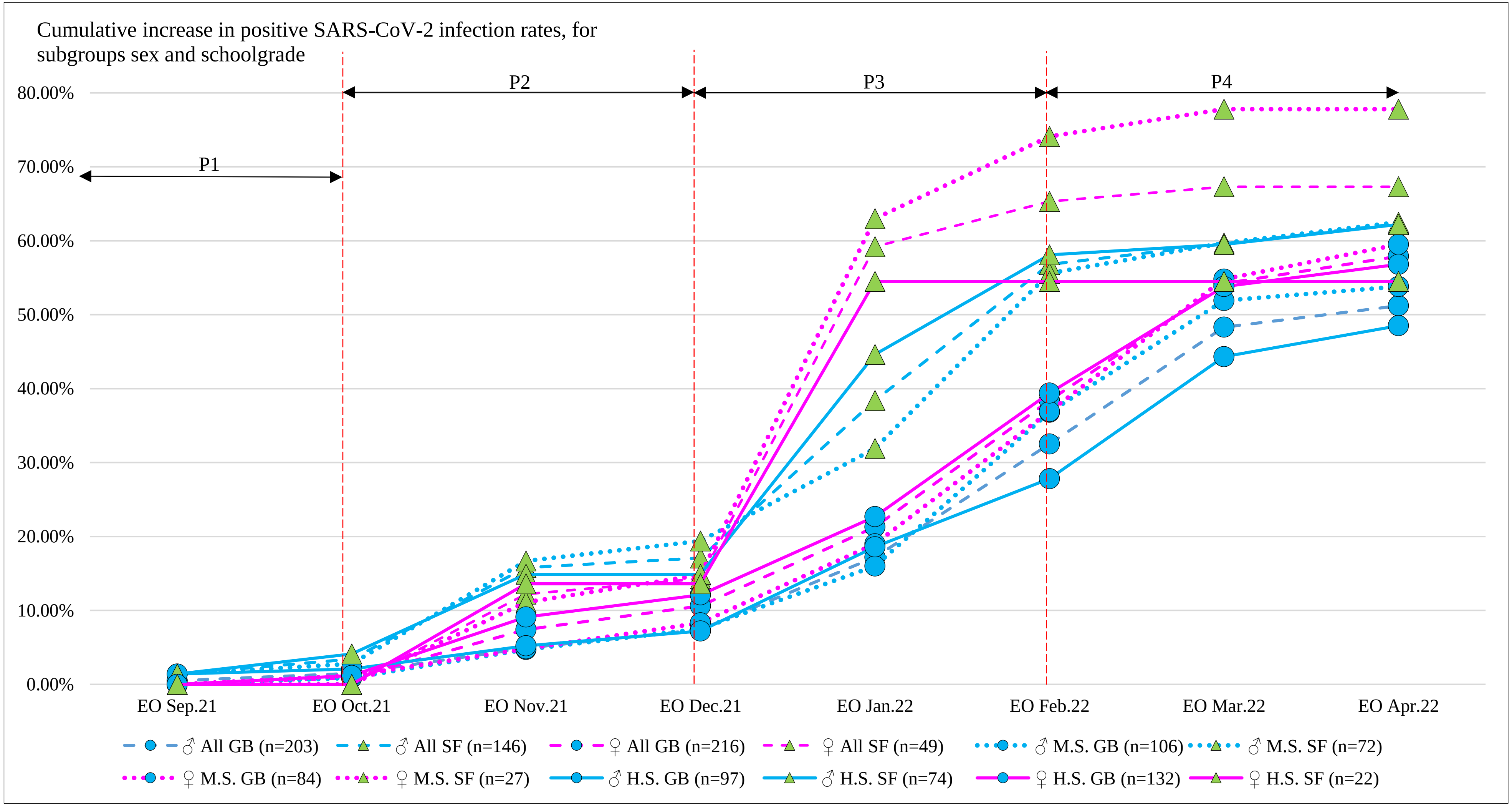


SARS-CoV-2 = severe acute respiratory syndrome coronavirus 2, P1 = period 1 (13. September to 31. October 2021), P1 = period 1 (September 13, 2021 to October 31, 2021) P2 = period 2 (November 1, 2021 to December 31, 2021), P3 = period 3 (January 1, 2022 to February 28, 2022), P4 = period 4 (March 1, 2022 to April 30, 2022), EO = End of, Sep.21 = September 2021, Oct. = October 2021, Nov.21 = November 2021, Dec.21 = December 2021, Jan.22 = January 2022, Feb.22 = February 2022, Mar.22 = March 2022, Apr.22 = April 2022, ♀ = Girls, ♂ = Boys, GB = students in school classes with a general school branch, SF = students in school classes with sport focus, M.S. = middle school (students aged 13.1 ± 1.3 years old), H.S. = 4-year high school (students aged 16.9 ± 1.2 years old), n = Study population.
